# Polygenic Health Index, General Health, Pleiotropy, Embryo Selection and Disease Risk

**DOI:** 10.1101/2022.06.15.22276102

**Authors:** Erik Widen, Louis Lello, Timothy G. Raben, Laurent C. A. M. Tellier, Stephen D. H. Hsu

## Abstract

We construct a polygenic health index as a weighted sum of polygenic risk scores for 20 major disease conditions, including, e.g., coronary artery disease, type 1 and 2 diabetes, schizophrenia, etc. Individual weights are determined by population-level estimates of impact on life expectancy. We validate this index in odds ratios and selection experiments using unrelated individuals and siblings (pairs and trios) from the UK Biobank. Individuals with higher index scores have decreased disease risk across almost all 20 diseases (no significant risk increases), and longer calculated life expectancy. When estimated Disability Adjusted Life Years (DALYs) are used as the performance metric, the gain from selection among 10 individuals (highest index score vs average) is found to be roughly 4 DALYs. We find no statistical evidence for antagonistic trade-offs in risk reduction across these diseases. Correlations between genetic disease risks are found to be mostly positive and generally mild. These results have important implications for public health and also for fundamental issues such as pleiotropy and genetic architecture of human disease conditions.

## 1 Introduction

Interest in polygenic risk scores (PRS) and the ability to estimate disease risks from genotypes has increased steadily over the past decade. A polygenic risk score maps an individual genotype to a score that reflects genetic risk for a particular disease; most PRS depend on hundreds or thousands of individual loci in the genome. As biobank data sets have grown larger, so have the performances and applicability of PRS. There are now a multitude of predictors that can assign estimated disease risks with an accuracy that has reached clinical utility. Disease conditions as diverse as coronary artery disease, breast cancer, and schizophrenia can be predicted with a useful accuracy from genetic information alone [1–21]. Typically, PRS are trained on and applied to a single disease but with many such risk predictions available it is natural to ask whether they could be combined into a general health index — a single number to describe the overall health of an individual. This question has already been explored in [22], where the authors created a composite PRS using a cox-hazard model, utilizing diseased participants of the UK Biobank (UKB). This composite PRS was found to predict longevity. The impact on longevity and individual disease burdens from individual variants has also been studied, using the Finish databank FinnGen [23].

In this paper, we construct a general health index by combining PRS for 20 diseases (**Table 1**), choosing the individual disease weights in an attempt to minimize the number of life years lost due to illness. We evaluate whether such a single number is a useful reflection of an individual’s various disease risks and their combined effect on estimated life years. If true, it could be a valuable tool for clinicians and patients to assess combined risks and genetic health predisposition. For a wide range of reasons, interpreting clinical risk based on genetic data can be difficult for both patients [24–29] and clinicians [30–32]. Combining PRS into a single metric can greatly simplify the process of evaluating genetic risk reports.

**Table 1:** Disease abbreviations and predictor AUCs. AUCs are listed with the standard deviations in parentheses, as computed by 30 bootstrap runs (see Supplementary Information).

| Abbr. | Disease | AUC | Abbr. | Disease | AUC | Abbr. | Disease | AUC |
| --- | --- | --- | --- | --- | --- | --- | --- | --- |
| AD | Alzheimer’s disease | .686<sub>(.004)</sub> | HA | heart attack | .580<sub>(.008)</sub> | Obes | obesity | .669<sub>(.002)</sub> |
| AFib | atrial fibrillation | .623<sub>(.004)</sub> | HCL | hypercholesterolemia | .616<sub>(.003)</sub> | PC | prostate cancer | .64<sub>(.02)</sub> |
| ASA | asthma | .626<sub>(.004)</sub> | HTN | hypertension | .635<sub>(.003)</sub> | SCZ | schizophrenia | .67<sub>(.03)</sub> |
| BC | breast cancer | .594<sub>(.008)</sub> | IBD | inflammatory bowel disease | .647<sub>(.003)</sub> | T1D | type I diabetes | .63<sub>(.02)</sub> |
| BCC | basal cell carcinoma | .62<sub>(.01)</sub> | IS | ischemic stroke | .541<sub>(.002)</sub> | T2D | type II diabetes | .616<sub>(.004)</sub> |
| CAD | coronary artery disease | .616<sub>(.005)</sub> | MDD | major depressive disorder | .534<sub>(.001)</sub> | TC | testicular cancer | .61<sub>(.04)</sub> |
| Gout | gout | .65<sub>(.01)</sub> | MM | malignant melanoma | .57<sub>(.02)</sub> |  |  |  |

Another prominent application of a general health index is to inform embryo selection in IVF cycles (in vitro fertilization). Embryos are routinely biopsied for aneuploidy and monogenetic disease tests. For cycles resulting in more than one euploid embryo (without any of the monogenetic disease variants), clinicians and prospective parents typically select which embryo to implant based on visually assigned embryo grades. With the advent of preimplantation polygenetic testing, a general health index could additionally be used to guide this choice and reduce the overall disease risk for the baby.

A priori, it is not given that such a health index would be useful. A common preliminary objection is that an index or single PRS, while reducing the disease for one disease, could inadvertently increase the risk for another [33, 34]. However, it has long been known that many diseases in reality tend to come together [35–46]. This could, at least for some broad categories of diseases, allow for useful indices. The specific concern raised for polygenic health indices has been the possibility of antagonistic pleiotropy, i.e., that a single gene may affect more than one disease risk simultaneously and in such a way that it decreases one disease risk while increasing another. If such pleiotropy were very common, there would not be much point of a genetically based health index.

In this paper, we examine both the underlying phenotypic comorbidities and genetic pleiotropy. The 20 studied diseases frequently occur together, sometimes with strong positive phenotypic correlation, while the genetic pleiotropy is usually small and slightly *positive*, or negligible. More importantly, we show in practice, using real genetic and health data, that the proposed health index can identify individuals at high or low risk for almost all diseases simultaneously. We observed individual disease risk reductions even beyond 40% (CAD, heart attack, diabetes type II) when selecting the highest index among five individuals, as compared to the general population. We further see no statistically significant evidence for inadvertent risk increments among any of the 20 diseases.

These conclusions are drawn from several experiments. We apply the constructed index to about 40,000 late-life individuals of European ancestry for whom both genotypes and medical history are known, using the UK biobank (UKB). Odds (prevalence) plots are shown for the most common diseases but the majority of the results are in form of selection experiments. The test data samples are grouped, using different group sizes in different experiments, and the sample with the highest health index is selected from each group. The selected individuals are then compared to the total test set to see the health differences in the medical history data, computing metrics like Relative Risk Reduction (RRR) and estimated gained life years. These experiments are repeated and confirmed with a very strong test of the genetic signal: selection among pairs (21,539) and trios (969) of genetic siblings. Siblings have both less genetic variation and typically share similar family environments, thus constituting an excellent test set. Finally, the underlying phenotypic and PRS dependencies are analyzed.

It is well-established that PRS are more accurate within a population ancestrally homogeneous and similar to the training population — however, generally a positive effect in one ancestry will persist in more distant ancestries. Research on this topic is ongoing and of high interest [7, 47–50]. The primary motivation for this paper is to investigate whether a composite genetic health index is reflective of general health in principle and we therefore focused on a single ancestry with maximum amount of data.

All analyses, except where otherwise indicated, are performed on self-reported white samples from the full UKB release (2021-04); these are almost exclusively of European ancestry. We set aside 39,913 samples as a pure test set, withheld from all training and hyperparameter tuning^1^. The PRS are constructed through a previously published LASSO-algorithm [7] trained on ∼ 200k-400k samples from the training portion of the same UKB data, except for the predictors for AD, IBD, IS, MDD, and SCZ. More details on the predictors can be found in the Supplementary Information.

## 2 Results

### 2.1 Overview of Methods

#### Polygenic health index

There are many ways to construct a polygenic health index from multiple PRS. Here we investigate the performance of a single linear combination of risk estimates, attempting to reduce lost life years. Let *l*_*d*_ be the estimated reduction in life expectancy for an individual having a disease *d* as compared to the general population, and let *ρ*_*d*_ be the lifetime risk in the general population of getting the disease. For the predicted risks *r*_*d*_, we define the health index to be

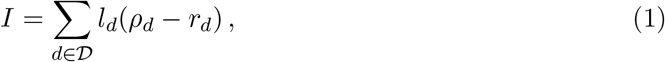

for a selected set of diseases *D* (this paper consistently uses the 20 diseases in **Table 1**). As such, a higher *I* should correspond to a healthier individual. As a proxy for ground truth in our test data set, we also define a case/control-based version, *I*^*c*^, which instead of the risk *r*_*d*_ uses the recorded case/control status *c*.^2^ We use this quantity as measure of the real world outcome value of the index. We note that the majority of our UKB test set is still alive (ageµ = 70, *σ* = 7 years) making *I*^*c*^ an imperfect measure of lifetime outcomes and skewed towards diseases with early onset. Still, since the mean age is not more than about one standard deviation (SD) from the average lifespan and the incomplete data masks cases as controls, rather than vice versa, we expect that a health index validated on an *I*^*c*^ using complete data (with perfect lifetime medical records and age of death) would have a better performance than what is measured in the UKB data.^3^

The index parameters *l*_*d*_ and *ρ*_*d*_ were taken from literature studies, using the average values if more than one source was used (see Supplementary Information).

#### Gaussian risk model

The health index definition (1) requires an estimated absolute (lifetime) risk *r* for each disease, modeled from the PRS as input. Depending on disease and predictor specifics, there are different possible choices for this modeling. A fairly general model, which works very well for sufficiently polygenic PRS (i.e., such that the Central Limit Theorem can be applied), models the PRS as drawn from a sum of two normal distributions with case/control status dependent means (*µ*_1_/*µ*_0_) and joint variance. The PRS probability distribution can then be written as

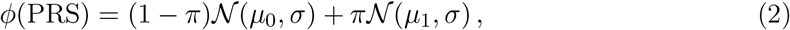

where *π* is the population prevalence and 𝒩 is the normal distribution. This leads to the Gaussian risk model

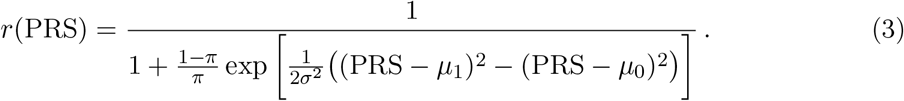

The case and control variances do not need to be equal in principle^4^ but in practice tend to be close in value (see Supplementary Information). We use estimates ofµ_0_,*µ*_1_, and *σ* based on the PRS in test set controls and cases.

#### Selection experiment from groups of unrelated individuals

To evaluate the performance of the health index, we created sets of groups and carried out selection experiments, i.e., we grouped together random individuals in the test set into groups of a specific size and than picked one individual from each group. In index selection experiments, we selected the individual with the highest index value. In PRS selection experiments we selected the individual with the lowest PRS (lowest risk) for a specific disease.

We created 40k random groups from the samples belonging to the intersection of all predictor test sets, such that no sample was used in any type of training nor hyperparameter tuning. Each sample was scored and assigned a raw and a sex-adjusted (see Supplementary Information) health index, as in equation (1). For each selection outcome, we calculated the relative risk reduction (RRR) for each individual disease and the index gain as measured in the case/control-based index *I*^*c*^, as compared to a completely random selection (i.e., the general population statistics):

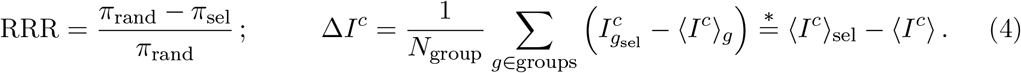

Here *g* sums over all *N*_group_ groups, 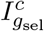 is the health index for the selected individual in group *g*, and ⟨·⟩ denotes the sample means, i.e., ⟨*I*^*c*^⟩_*g*_ is the average health index value in group *g*, ⟨*I*^*c*^⟩ _sel_ is the average among all selected individuals, and ⟨*I*^*c*^⟩ is the average in the total test set. The index gain Δ*I*^*c*^ can be viewed either as the average index difference between the selected individual and its group average or as the difference between the average selected index and the general population average (∗ holds for constant group size). Note here that we are using the case/control status based index, *I*^*c*^, as evaluation metric which does not use any genetic information but only individual lifetime^5^ disease status, together with the population based lifespan impact and lifetime risk estimates. The full selection experiment procedure is illustrated in **Figure 1**.

**Figure 1:**
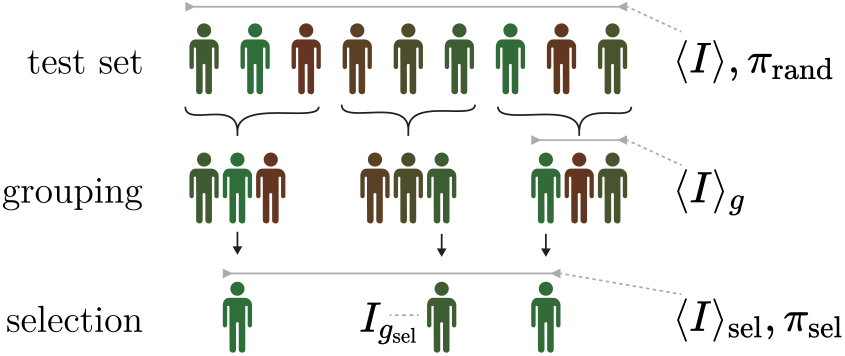
The selection experiments. The test set is scored with the health index *I* or a single PRS and is randomly divided into groups of equal size. The individual with the best score in each group is selected and the health status among the selected are then compared with the general test set. The symbols in equation (4) refer to indicated subsets.

We repeated all selection experiments 25 times to get a bootstrap estimate of the errors, reusing the same samples but grouping them into different groups. Thus, these are underestimates neglecting the additional variance that would come from also using other samples, while the group constellations are practically unique.

For the three sex specific diseases (breast, prostate and testicular cancer), we compared only the subsets with the relevant sex of the selected and random sets when calculating the RRR and index gain.

#### Genetic sibling selection

The selection experiments on unrelated individuals provide good metrics for how the health index performs in the general population. A much stronger test is to repeat the same experiments using real world siblings, sharing half their genetic material. Accurate prediction within siblings is challenged both by this reduced genetic variance and by more similar environments; it is thus a rigorous test of genetic prediction performance.

We repeated the selection experiments for 21,539 pairs and 969 trios of genetic siblings. Since the sibling data cannot be re-grouped as in the unrelated selection experiments, we opted to not use bootstrap errors but instead calculate the theoretical 95% confidence interval for the prevalence among the selected siblings, based on the Wilson score interval. It was translated to the RRR metric through equation (4), keeping the population prevalence *π*_rand_ fixed. We did not estimate the errors for the index gain metric when selecting among genetic siblings.

### 2.2 Selection experiment using groups of unrelated individuals

We report the overall index gain (Δ*I*^*c*^ from eq. (4)) from the selection experiments on unrelated individuals in **Figure 2**. It documents a well-established and consistent gain that increases with group size, maintaining a positive increment even when selecting among more than 10 people. The health index distribution is non-Gaussian with standard deviation (SD) of 1.56 estimated life years and with a skewness of −0.49. The difference between the mean health index values for the top and bottom 5% of the index *I* was 5.10 predicted life years. The corresponding difference between these groups was 3.49 years when measured with the case/control based index *I*^*c*^ (a smaller difference is to be expected due to the incomplete case/control data). Despite different methods and disease sets, we note the connection to [22] which reported similar values in lost life years per SD and difference between top and bottom 5% of composite PRS. In **Figure 3**, the selection experiment result at the group size of five is broken down into the RRR and the component-wise index gain for each disease, allowing a more fine-grained view of the performance. Strikingly, the RRR graph is overwhelmingly positive thus demonstrating compelling evidence that selected individuals with higher health index score have lower incidence for almost all diseases at the same time. 15 out of the 20 disease have statistically significant positive RRR, reaching over 40% for the most reduced disease risks (CAD, HA, T2D), whereas none is significantly negative or even has a negative central value. It is important to note that although the weights *l*_*d*_ matter for how the index is constructed and thus for whom is selected, they have no direct impact on the RRR metric itself — only the actual disease status is measured. As such, the RRR plot is a true measurement of the reduced disease incidence. In contrast, the right plot in **Figure 3** of the index gain Δ*I*^*c*^ involves the weights both in selection and in evaluation. Using the weights based on estimated lost life years, we get a disease-by-disease breakdown of the index gain. Again, there is a statistically significant positive contribution from almost all diseases with obesity, type II diabetes, major depressive disorder and CAD as the strongest contributors.

**Figure 2:**
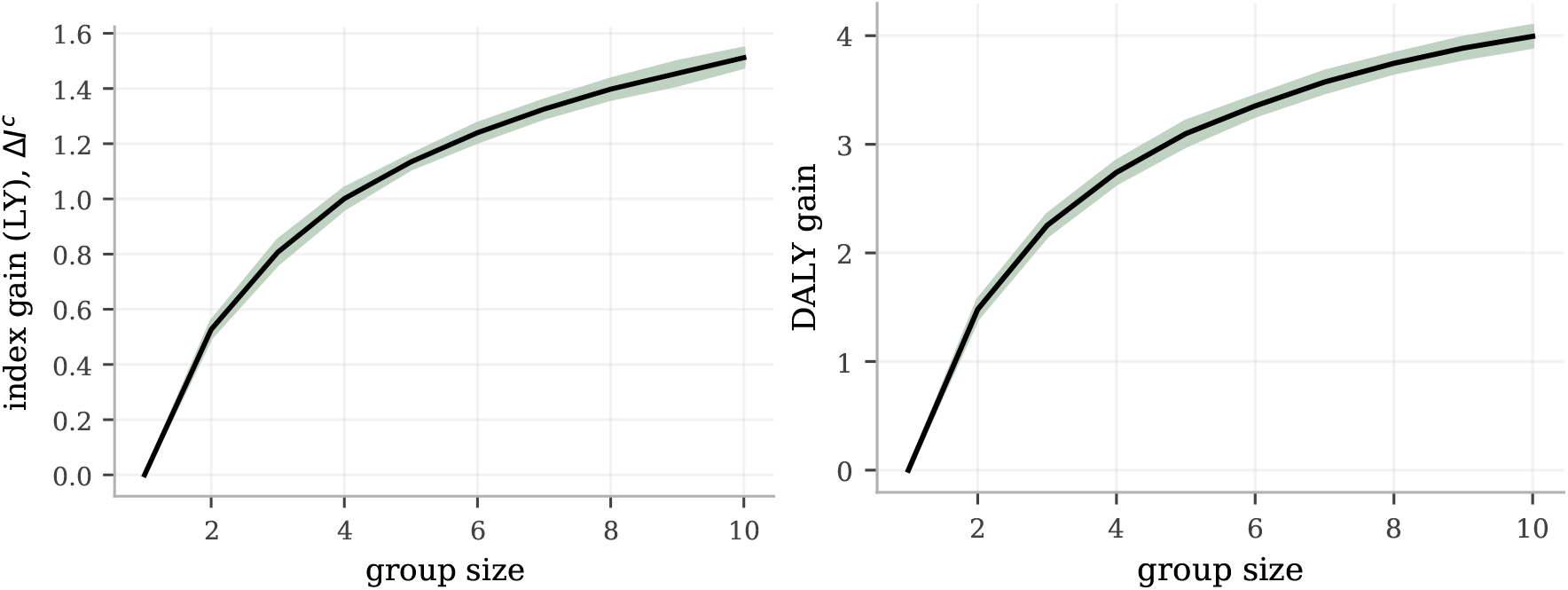
The estimated gain from index selection is a clearly positive function of group size, both using disease weights as defined by population estimates of lost life years or using disease weights based on disabilty-adjusted life years (DALY). **Left:** The index gain, as measured as the average health index difference between selected and random individuals (Δ*I*^*c*^ in eq. (4)), is growing monotonically with group size and with a continued clear positive derivative at group sizes of 10. Notably, there is a strongly significant gain for all group sizes, even at a group size of 2. The error band is a 95% C.I. as computed by 25 experiments with independent selection groupings. **Right:** While still selecting on the same index (1), we evaluated it on a case/control status metric using DALY-weights, taking quality of life into account. Again, there is a clear and steady gain, with the gain at a group size at 10 reaching about 4 years. The error band is a 95% C.I. as computed by 25 experiments with independent selection groupings.

**Figure 3:**
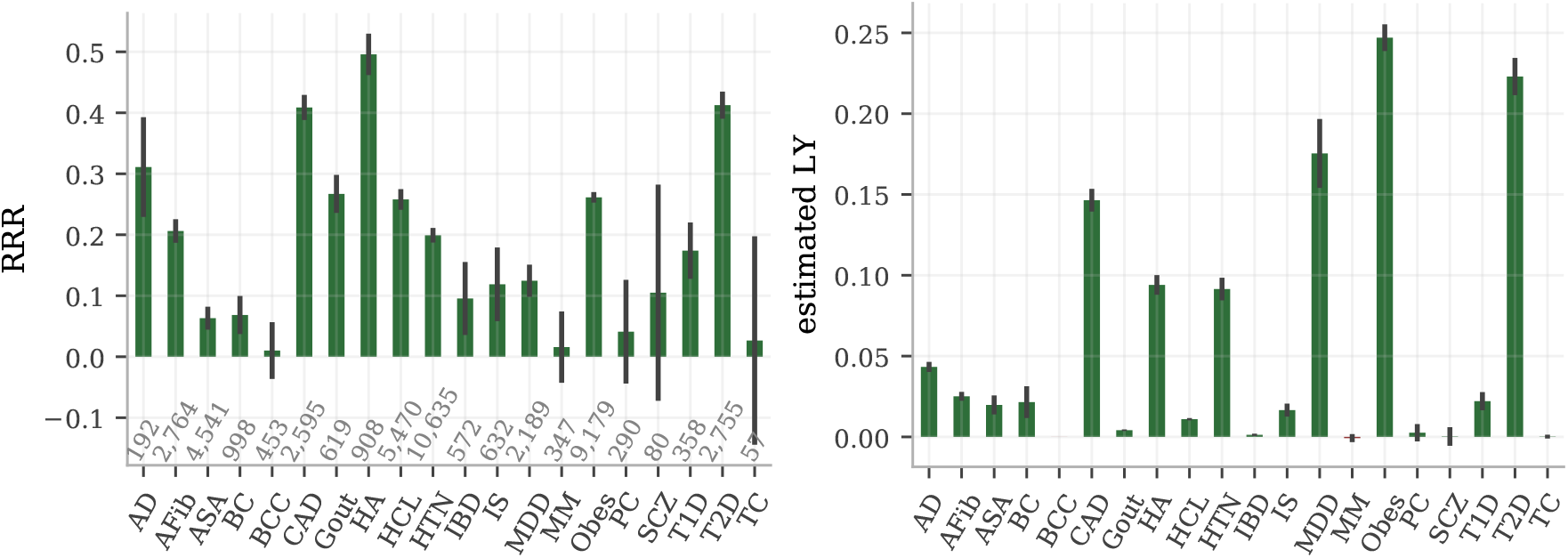
Selecting on health index among 5 randomly grouped individuals reduces simultaneously the risk of almost all the studied diseases. **Left:** The RRR among the selected individuals as compared to random selection is dominantly positive, ranging from a few risks reductions statistically consistent with zero up to more than 40%. No disease risk is demonstrably increased. The case numbers for each disease are printed just above the x-axis and the error bars are 95% C.I. estimates from 25 repeated experiments with different selection groupings. **Right:** The estimated index gain for each of the index components (diseases), i.e., the disease component breakdown of equation (4), also shows non-negative gains across the board with most component gains being statistically significant. The unit on the y-axis is estimated life years (LY), as is the unit of *I*^*c*^. This index is primarily driven by CAD, heart attack, hypertension, major depressive disorder, obesity and type II diabetes, due to their combinations of strong impacts *l*_*d*_ and high population prevalence.

The average component gains in **Figure 3** depend both on the quality of the individual PRS, the weights *l*_*d*_ and the test set prevalences. For example, the AD predictor has a much stronger individual performance than MDD (AUC ∼ .69 vs ∼ .53) while MDD has stronger weights than AD in the index (*l*_MDD_*/l*_AD_ ≈ 1.6). The index achieves a RRR of about 31% for AD and 12% for MDD, with the individual PRS-performance having a larger impact on the RRR metric. Meanwhile, MDD has about 4 times the AD contribution to the index gain, largely due to it being about 10 times more prevalent in the test set. Naturally, common diseases contribute more to the average index difference than rare ones. Both AD and MDD have some strong comorbidities and milder PRS-correlations with other diseases; this is discussed further in section 2.4. See also the Supplementary Information for a deeper discussion of the test set prevalences and their influence on the quantitative results.

The RRR and index gain metrics offer complementary information of the potential bene-fits: the RRR captures how much the risk can be reduced simultaneously, while the index gain translates this into estimates of the corresponding life years gained on average. All selection experiments selected on the index in equation (1), using lost life years *l*_*d*_ as weights. A common alternative for assigning relative importance to diseases is the unit Disability Adjusted Life Years (DALY). While still *selecting on our index* (1), we make contact to the existing DALY-literature by *evaluating the index gain* using a DALY-scale to the right in **Figure 2**. The weights in the evaluating index difference Δ*I*^*c*^ were computed as population level DALY-oefficients *l*_*d*_ + *q*_*d*_Δ*y*_*d*_, where *q*_*d*_ is a disability factor between 0 and 1 and Δ*y* is the number of years between average age of onset and average age of death. The individuals selected from groups of size 10 had an increase of 4 DALY as compared to randomly selected individuals. This magnitude scale comports with previous studies [23].

The index tries to minimize the risk for several diseases simultaneously. In **Figure 4** we demonstrate how all the RRR from index selection compare to the RRR when selecting directly on the individual disease PRS, i.e., how much the index retains of the maximal risk reduction you would achieve if you focused on reducing a single disease. The direct PRS-selection tend — as naively expected — to reduce the specific diseases risk more than the index, especially for those diseases with very small weights (BCC, IBD). Yet, there are several examples where the index actually matches or even surpasses the direct PRS performance, most notably HA (probably because the strong/large comorbidity with CAD, HTN and obesity).

**Figure 4:**
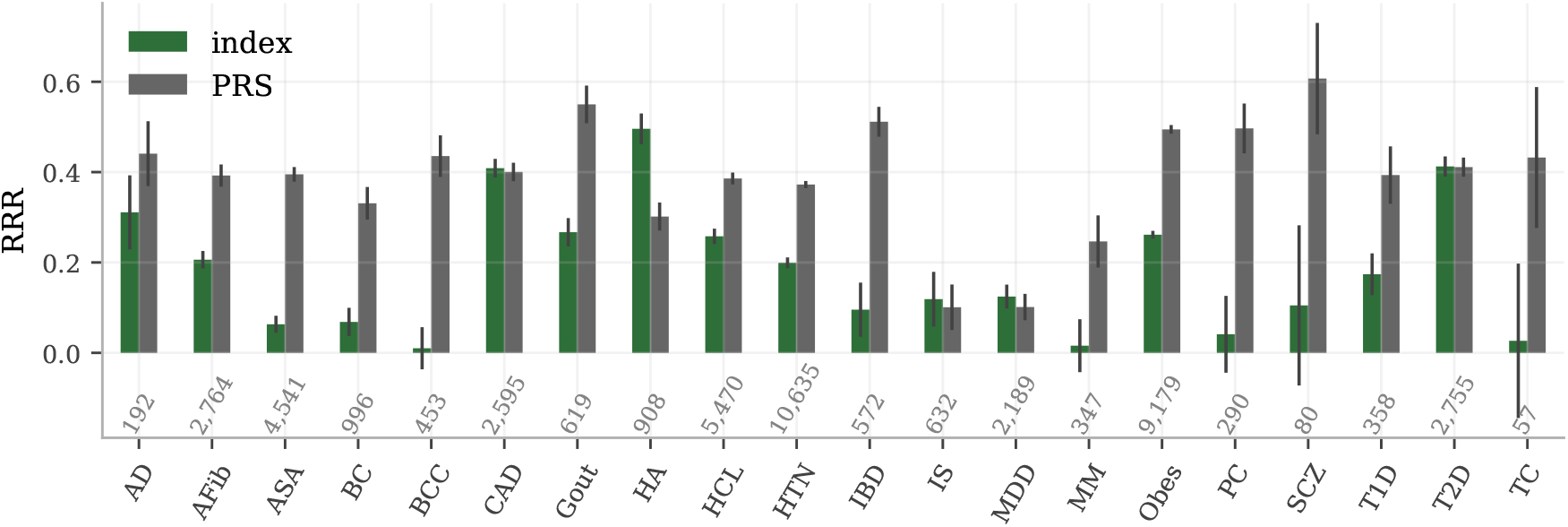
RRR comparison between selection on index and selecting on individual disease PRS. The individual disease RRR obtained by index selection contrasted with selection directly on the individual PRS, using a group size of 5. The case numbers in the test set for each disease are shown above the x-axis and the error bars are 95% C.I. as computed by 25 independent experiment runs.

The PRS-comparison in **Figure 4** is a cross-section of the results at a group size of 5. The patterns are however consistent across all tested sizes, as seen in **Figure 5**. The index reduces the risk of both T2D and CAD by about 50% at group size 10, consistently matching both the individual PRS-performances simultaneously. The consistent difference between PRS and index selection are also shown for Alzheimer’s disease and obesity.

**Figure 5:**
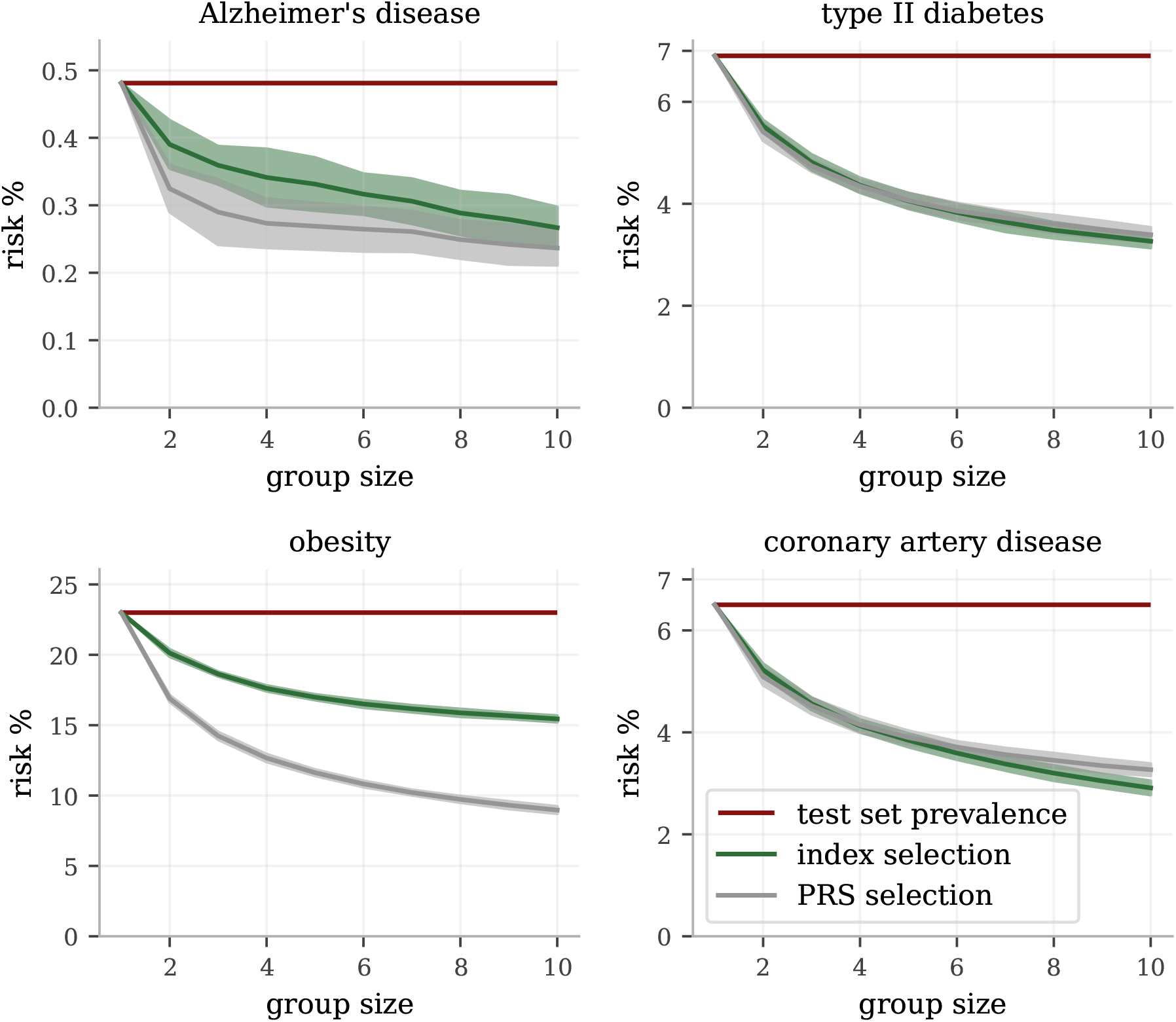
The disease risk reduction from index and PRS selection for different group sizes. The relative performance between index selection and PRS for individual diseases varies, as seen in **Figure 4**. Here shown as functions of the group size, we see the strongest performance step between having no selection (group size 1) and selecting between between two and also the continued, but less dramatic, benefits with larger group sizes. Notably, for the chosen examples type II diabetes and CAD, the full health index consistently perform as well as selecting directly on the specific PRS, showing no reduced effects on these disease from taking all the other into account. The index performance for Alzheimer’s disease and obesity, while not achieving the full risk reduction of their corresponding PRS, retain significant risk reductions for all group sizes. The error bars represent estimated 95% C.I. as computed by 25 selection experiments using different selection groupings.

For the most prevalent diseases (ASA, HCL, HTN and obesity), we also provide prevalence-per-index quantile plots (odds ratio plots if divided by the general prevalence) in **Figure 6**; the less prevalent diseases did not have enough cases for such high resolution. The top 4 percentiles have about half the risk of the bottom 4 percentiles to have either of hypercholesterolemia, hypertension, and obesity, while the risk reducing trend for asthma is less dramatic.

**Figure 6:**
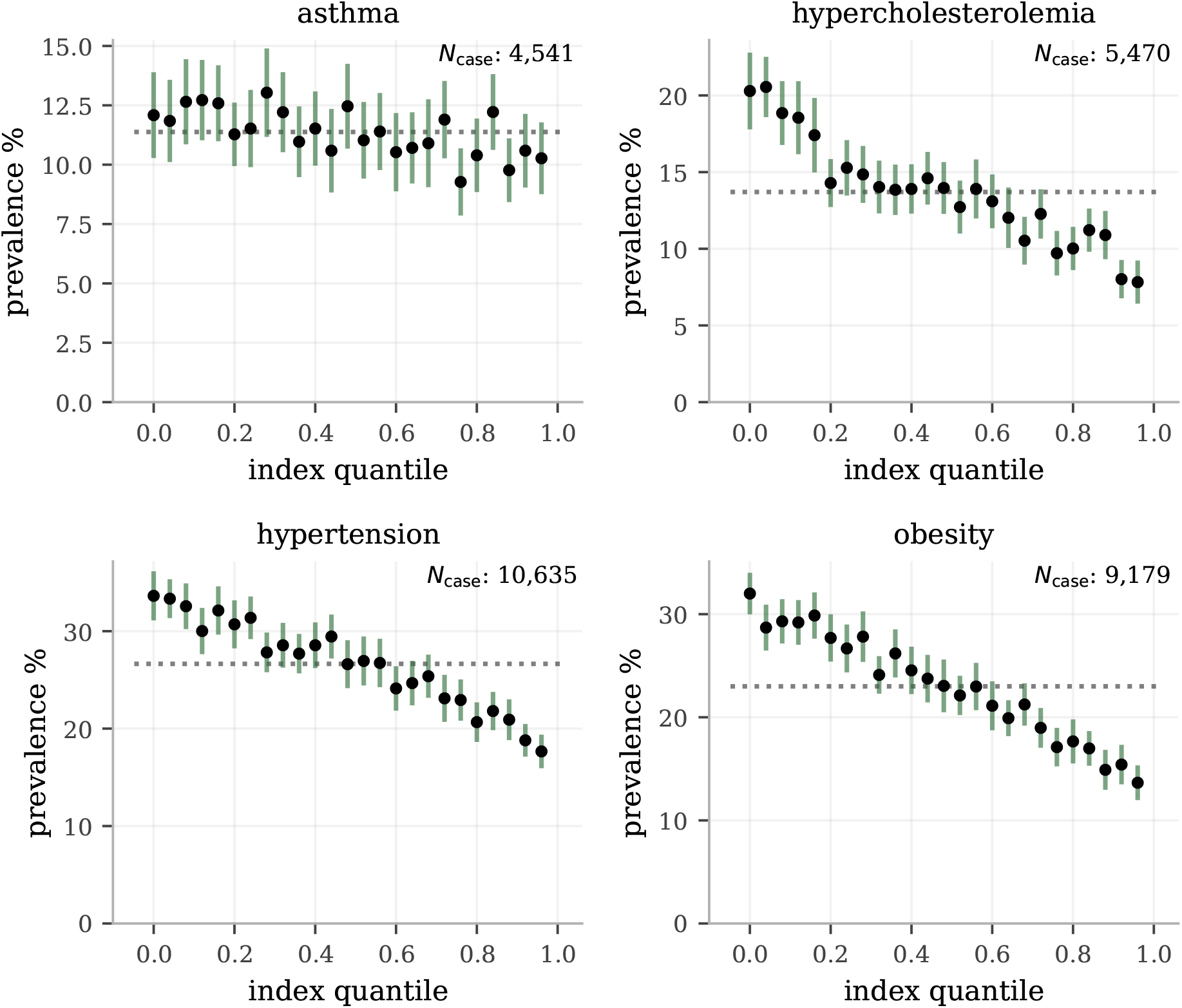
Prevalence in health index quantile bins for the most common diseases. We binned the test set according the health index into 25 equally distributed quantiles and plot the prevalence within each bin for the most prevalent diseases (allowing enough cases for the bin resolution to be meaningful). The general population prevalences are plotted as dotted reference lines (dividing with this number would give odds ratio plots) and the y-axis start at 0 to give a visual representation of the (odds) scales. For the intermediately risk reduced diseases (according to RRR **Figure 3**) hypercholesterolemia, hypertension and obesity, there is a clear and systematic risk relationship across the entire range of the health index. For asthma, there is only a weak, detectable trend for the center values consistent with its existing but smaller RRR. The error bars are 95% C.I. estimates obtained through 100-fold bootstrap calculations of the prevalence within each bin (no re-binning was done).

### 2.3 Genetic sibling pairs and trios

The primary results for the selection experiment on pairs of siblings is shown in **Figure 7**, broken down into RRR and component index gain for each disease. The same graphs also include as reference the results from the selection among unrelated samples at group size 2. The sibling with the largest health index was selected from each of the 21,539 sibling pairs; no bootstrap was carried out. Instead the RRR error bars for the genetic siblings are theoretical 95% confidence intervals using the Wilson score interval for the prevalences among the selected siblings. They are generally larger than the corresponding error bars for the group size 2 bootstrap experiment. The limited data, for the rarest diseases in particular, decrease the certainty and result in the large error bars. Yet, we conclude from **Figure 7** that even in the most challenging task of minimizing the disease risk among only two genetic siblings the index provides a simultaneous and verifiable reduction of many diseases, while others are left inconclusive in this data set. Among the 20 studied diseases, there is no example of verified increased disease risk. Similarly, the estimated index gain is non-negative for all disease components and sum up to a significant gain also among pairs of genetic siblings.^6^

**Figure 7:**
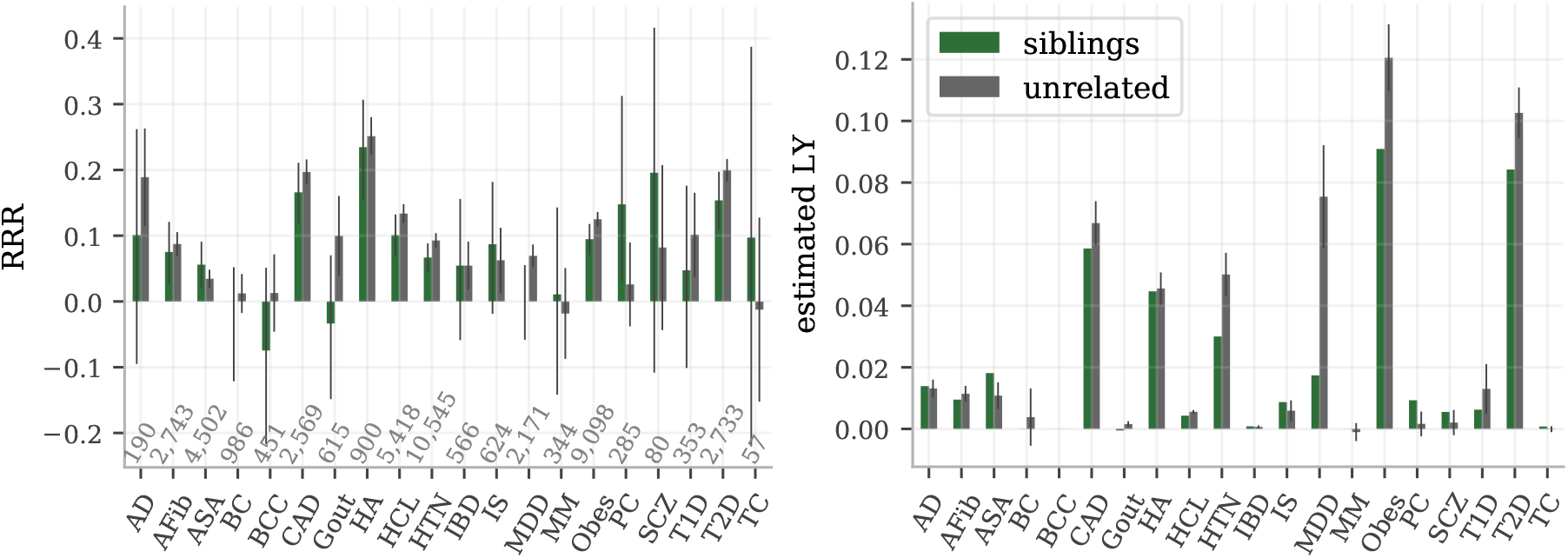
Index selection between 22,667 pairs of genetic siblings retain the overall benefits. In both figures, selection experiments among pairs of genetic siblings are compared to selection among pairs of unrelated individuals. The index performances are qualitatively very similar despite that siblings share half their genomes and have more similar environments. As expected, we do see a general performance attenuation among siblings, but also a few exceptions. **Left:** The RRR for each disease. The error bars for siblings are theoretical 95% C.I. using Wilson score interval for the prevalences among the selected siblings. The error bars for the selection among the unrelated pairs are again estimated 95% C.I. from 25 separate runs. The case numbers are shown above the x-axis. **Right:** The component-wise index gain for the selections among pairs of siblings and among pairs of unrelated individuals. The sibling results are presented without error bars since no theoretic uncertainty was calculated; statistical significance is therefore not established from this data. The error bars for the selection among unrelated individuals are 95% C.I. from 25 separate runs.

The index selection experiment result on the 969 trios had to the most part large uncertainties due the smallness of the data set and low case counts. Only two disease RRR reached statistical significance, according to the theoretical RRR confidence intervals. Hypercholesterolemia and obesity were confirmed with positive RRR, while hypertension and type II diabetes bordered to positive significance. No disease was confirmed to have negative RRR. The full RRR and index gain plots for trios are to be found in the Supplementary Information.

### 2.4 Characterization of phenotypic and genetic dependencies

The simultaneous disease risk reduction demonstrated for the index selection is bounded by potential disease dependencies, i.e., if two or more diseases tend to occur together (comorbidity) or are mutually exclusive. A commonly raised concern for PRS, and even more so for a composite health index, is the risk of antagonistic pleiotropy, i.e., that the same gene simultaneously increases the risk for one disease while decreasing the risk for another. Such a situation (or any cause of negatively correlated disease incidence) would impede simultaneous risk reduction. We examined this question for the 20 chosen diseases within our test set both on a genetic and phenotypic level. The result is presented in **Figure 8** through three quantities for each pair of diseases: the correlation between the PRS, the ratio between observed and expected comorbidity (called the *χ*^2^ ratio), and the p-value of a *χ*^2^ independence test (see figure caption for the details of the quantity visualization). The high information density in the plot requires some explanation but allows for quick comparison between all three quantities, both for individual pairs and for the disease set as a whole.

**Figure 8:**
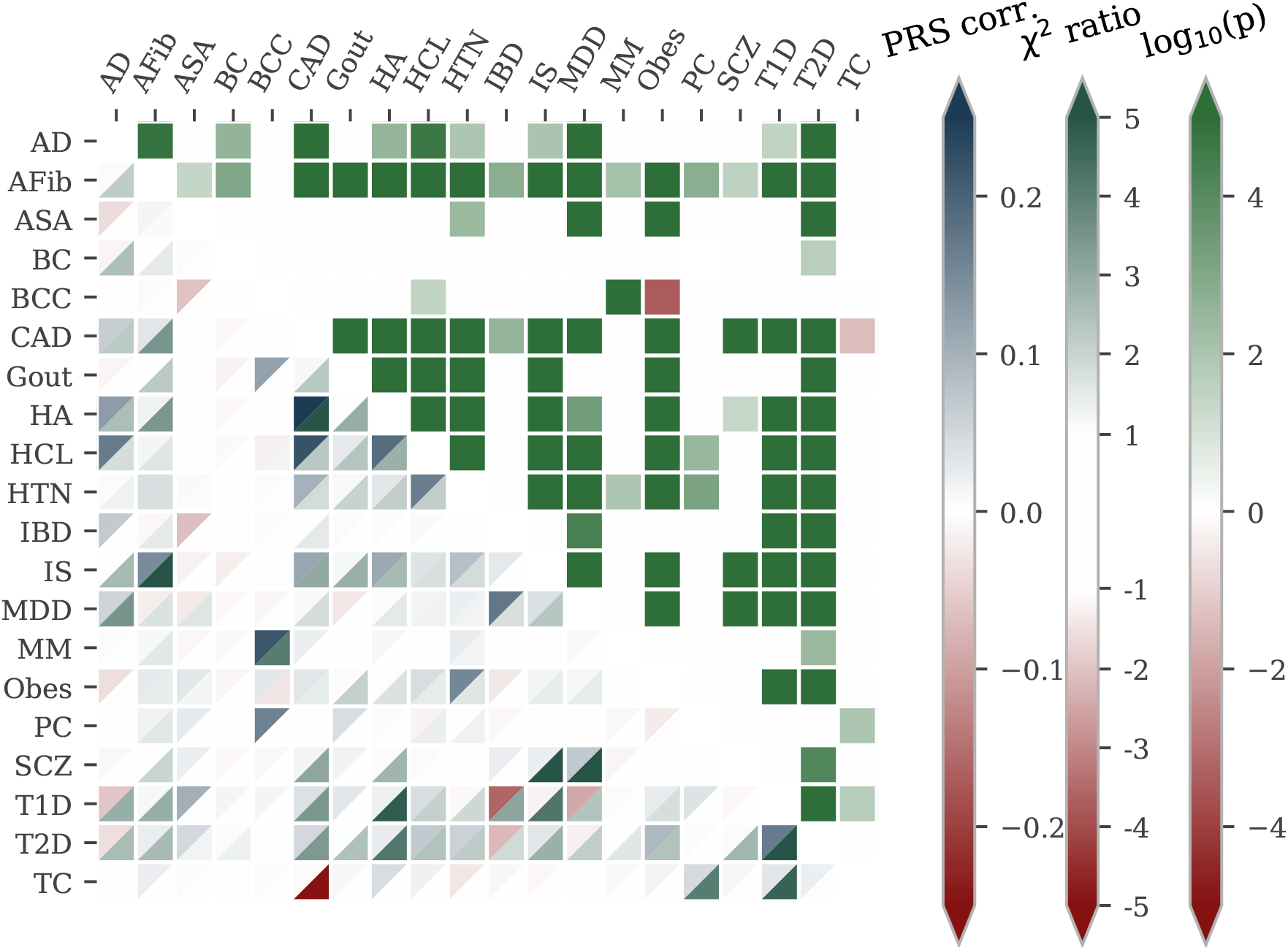
Phenotype dependencies and PRS correlation comparisons. This figure visualizes three different quantities for each pair of disease: the PRS correlation, a comorbidity metric, and a *χ*^2^ independence test p-value. Each tile **below the diagonal** is split into two halves where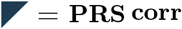. is the correlation between the two diseases’ PRS, i.e., the genetic correlations as inferred by the predictors. The other half, 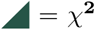 **ratio**, is a metric of the actual disease comorbidity: how many more times is disease coincidence observed compared to what would be expected if the diseases were completely independent, where a positive (negative) sign indicates higher (lower) comorbid frequency (this is based on the ratio between the observed and expected case-case cell in a *χ*^2^-test contingency table, hence referred to as the *χ*^2^ ratio). The squares,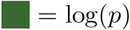, **above the diagonal** indicate the statistical significance of the dependence: the (signed) logarithm of the p-value in a *χ*^2^-test. The sign is positive (negative) for more (less) frequent coincidence. Both the p-value and the *χ*^2^ ratios are masked for disease pairs without statistically significant (*p* = .05) dependence. For example, the deep green square above the diagonal at (CAD, HCL) indicates that the CAD-hypercholesterolemia comorbidity is highly significant (we can reject phenotype independence at p-value *<* 10^−4^). Below the diagonal, we see for the same disease pair that the lower triangle is gently blue-green, i.e., case coincidence for CAD-hypercholesterolemia is about 2.3 times more common than random chance. Lastly, the upper triangle is dark blue meaning that the PRS correlation between CAD and hypercholesterolemia is among the very strongest, at about 0.22. Overall, we see that most disease pairs have statistically significant comorbidity with 1-2 times more coincidence than chance, and that their PRS are not, or slightly positively, correlated. This phenotypic and genetic background not only allows but facilitates the construction of a useful health index. The most prominent outliers are discussed in the main text.

Contrary to the concern about strong impacts of antagonistic pleiotropy, we find that the disease incidences typically are pairwise dependent and overwhelmingly occur together. The predominantly solid green squares above the diagonal confirm that most of the disease pairs have comorbitities of statistical significance, in line with longstanding results such as coincidence of CAD and hypercholesterolemia. This makes a health index not only possible but an almost natural concept. The *χ*^2^ ratio, 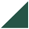-triangles below the diagonal, demonstrates the magnitude of the comorbidities, for example the very strong coincidences of (HA, CAD), (SCZ, MDD) and (T2D, T1D), and the moderate (HTN, AFib), (HTN, CAD) and (HCL, HA). The PRS correlations (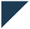 -triangles) are relatively small in magnitude and in general agreement with the phenotypic coincidences. As such, most PRS are relatively uncorrelated. Some notable exceptions are (HCL, CAD) and (MM, BCC). Just as the large amount of comorbidity facilitates the simultaneous positive RRRs, there are also some explanations for the lesser reductions here. The mutually exclusive tendency of (TC, CAD) complicates simultaneous risk reduction on a phenotypic level^7^. This is in accordance with **Figure 4**, where the RRR of TC is much stronger in PRS selection than index selection. The only examples of PRS level conflicts are the moderate anti-correlations between (T1D, IBD), (T1D, MDD) and (T2D, IBD), and the milder (BCC, ASA) and (IBD, ASA) anti-correlations, despite that these disease pairs are independent or have mild comorbidities. The combined index weights for ASA, T1D and T2D dwarf the impact of IBD on the index while BCC has no weight and is almost independent from everything else but MM (which is also independent from everything else). This contributes to the stronger RRR of PRS selection for ASA, BCC, IBD, and MM as compared to index selection.

## 3 Discussion

It is commonly believed that some individuals, in part due to genetic causes, enjoy robust good health, while others are sickly and prone to disease. Longevity is said to run in families. With modern genomic methods we can test the scientific veracity of these ideas. By combining Polygenic Risk Scores (PRS) across the most impactful disease conditions, we can build a composite predictor of overall health. The specific implementation studied in this paper used lifespan impact of each disease condition as the weighting factor in the index. We could then test whether this index predicts individual disease risks, as well as estimated longevity or disability adjusted life years.

Specifically, we validated this index in selection experiments using unrelated individuals and sibling pairs and trios from the UK Biobank. Individuals with higher index scores have decreased risk of individual diseases across almost all 20 diseases, with no significant risk increases, and longer calculated life expectancy. When Disability Adjusted Life Years (DALYs) were used as the performance metric, the gain from genetic selection (highest index score vs average) among 10 individuals was found to be roughly 4 DALYs, and among 5 individuals was found to be 3 DALYs.

We found no statistical evidence for strong antagonistic trade-offs in risk reduction across these diseases. Correlations between disease risks are found to be mostly positive, and generally mild. This supports the folk notion of a general factor which characterizes overall health, sometimes described as synergistic pleiotropy. These results have important implications for public health and also for fundamental biological questions such as genetic architecture of human disease conditions.

The concept of pleiotropy was formulated before the notion of high dimensional spaces of genetic variation became familiar. The conventional logic is that, because a single gene can affect many different complex traits, it must be the case that different complex traits, such as disease risks, are themselves correlated, perhaps antagonistically (e.g., due to balancing selection, or for some deeper biochemical reason). This would entail specific trade-offs, hypothetically: an individual with low diabetes risk might necessarily have higher cancer risk, etc. However, results from the modern era of GWAS and machine learning on large data sets show that the number of genetic loci which control a specific complex trait is typically in the thousands, and that these SNP sets are largely disjoint for different traits or disease risks [54]. The fact that most of the variance can be disjoint across different complex traits is a manifestation of high dimensionality. In this work we focus on *sparse* algorithms applied to array data which leaves open the possibility that there is are underlying causal loci that could still be correlated. However, the relatively small genetic correlations observed here leave this as an unlikely scenario.

In an earlier paper [54], we looked at the extent to which SNPs used in polygenic predictors of risk are correlated across pairs of disease conditions. Here we went further and investigated pairwise correlations between each of 20 major disease PRS. The results, as summarized in **Figure 8**, can be expressed in words as: most correlations are modest^8^, and tend to be positive rather than negative (antagonistic). We also concluded, on a phenotypic level, that the 20 diseases tend to have positive significant pairwise comorbidity.

We focused this paper on index performance in a single cohort, and carried out cross-cohort analyses in other populations. With increased data availability, these cross-cohort analyses will be expanded in scope.

## Data Availability

Access to the UK Biobank resource is available via application (http://www.ukbiobank.ac.uk).

## 4 Acknowledgements

Computational resources provided by the Michigan State University High-Performance Computing Center. The authors acknowledge acquisition of data sets via UK Biobank Main Application 15326.

## Footnotes

1 See the Supplementary Information for details on the test set.

2 Since there is a very large overlap between the case definitions we used for CAD and HA, we choose to exclude HA from the case/control based index *I*^*c*^. Otherwise HA would practically be double-counted in the performance evaluation.

3 The Supplementary Information contains more characterization of the test data.

4 Unequal variances can lead to unrealistic behavior in the tails.

5 see Supplementary Information for details

6 The mean values for BCC and Gout are negative but much smaller in magnitude than the uncertainty.

7 We are not aware of any research supporting this finding in other data sets. On the contrary, there are several examples of either inconclusive results or increased comorbidity of CAD among patients having undergone chemotherapy in TC treatment [51–53]. With our barely significant finding and small TC statistics, we view this result as peculiarity of the test set rather than a general epidemiological result.

8 Modest correlation is consistent with mostly but not entirely disjoint variance in the two PRS.

## References

1. Lewis, C. M. & Vassos, E. Polygenic risk scores: from research tools to clinical instruments. Genome medicine 12, 1–11 (2020) (cit. on p. 1).

2. Lewis, A. C. & Green, R. C. Polygenic risk scores: from research tools to clinical instruments. Genome medicine 13, 14 (2021) (cit. on p. 1).

3. Richardson, T. G., Harrison, S., Hemani, G. & Smith, G. D. An atlas of polygenic risk score associations to highlight putative causal relationships across the human phenome. eLife 8, e43657 (2019) (cit. on p. 1).

4. Wray, N. R. et al. From Basic Science to Clinical Application of Polygenic Risk Scores: A Primer. JAMA Psychiatry. issn: 2168-622X. https://doi.org/10.1001/jamapsychiatry2020.3049. (Sept. 2020) (cit. on p. 1).

5. Torkamani, A., Wineinger, N. E. & Topol, E. J. The personal and clinical utility of polygenic risk scores. Nature Reviews Genetics 19, 581 (2018) (cit. on p. 1).

6. Lello, L., Raben, T. G., Yong, S. Y., Tellier, L. C. & Hsu, S. D. H. Genomic prediction of 16 complex disease risks including heart attack, diabetes, breast and prostate cancer. Sci Rep 9. [PMC6814833], 1–16 (2019) (cit. on p. 1).

7. Widen, E., Raben, T. G., Lello, L. & Hsu, S. D. H. Machine Learning Prediction of Biomarkers from SNPs and of Disease Risk from Biomarkers in the UK Biobank. Genes 12. issn: 2073-4425. https://www.mdpi.com/2073-4425/12/7/991 (2021) (cit. on pp. 1, 3).

8. Wray, N. R., Yang, J., Goddard, M. E. & Visscher, P. M. The genetic interpretation of area under the ROC curve in genomic profiling. PLoS genetics 6 (2010) (cit. on p. 1).

9. Veenstra, D. L., Roth, J. A., Garrison Jr, L. P., Ramsey, S. D. & Burke, W. A formal risk-benefit framework for genomic tests: facilitating the appropriate translation of genomics into clinical practice. Genetics in Medicine 12, 686 (2010) (cit. on p. 1).

10. Amir, E., Freedman, O. C., Seruga, B. & Evans, D. G. Assessing women at high risk of breast cancer: a review of risk assessment models. JNCI: Journal of the National Cancer Institute 102. Oxford University Press, 680–691 (2010) (cit. on p. 1).

11. Euesden, J., Lewis, C. M. & O’reilly, P. F. PRSice: polygenic risk score software. Bioinformatics 31, 1466–1468 (2014) (cit. on p. 1).

12. Abraham, G. et al. Accurate and Robust Genomic Prediction of Celiac Disease Using Statistical Learning. PLOS Genetics 10, 1–15. https://doi.org/10.1371/journal.pgen.1004137 (Feb. 2014) (cit. on p. 1).

13. Priest, J. R. & Ashley, E. A. Genomics in clinical practice 2014 (cit. on p. 1).

14. Jacob, H. J. et al. Genomics in clinical practice: lessons from the front lines. Science translational medicine 5, 194cm5–194cm5 (2013) (cit. on p. 1).

15. Shieh, Y. et al. Breast cancer risk prediction using a clinical risk model and polygenic risk score. Breast Cancer Research and Treatment 159, 513–525 (2016) (cit. on p. 1).

16. Bowdin, S. et al. Recommendations for the integration of genomics into clinical practice. Genetics in Medicine 18, 1075 (2016) (cit. on p. 1).

17. Chatterjee, N., Shi, J. & García-Closas, M. Developing and evaluating polygenic risk prediction models for stratified disease prevention. Nature Reviews Genetics 17, 392 (2016) (cit. on p. 1).

18. Liu, L. & Kiryluk, K. Genome-wide polygenic risk predictors for kidney disease. Nature Reviews Nephrology 14, 723–724 (2018) (cit. on p. 1).

19. Nelson, H. D., Pappas, M., Cantor, A., Haney, E. & Holmes, R. Risk assessment, genetic counseling, and genetic testing for BRCA-related cancer in women: updated evidence report and systematic review for the US Preventive Services Task Force. Jama 322, 666–685 (2019) (cit. on p. 1).

20. Kulm, S., Marderstein, A., Mezey, J. & Elemento, O. A systematic framework for assessing the clinical impact of polygenic risk scores. medRxiv, 2020–04 (2021) (cit. on p. 1).

21. Wray, N. R. et al. From basic science to clinical application of polygenic risk scores: a primer. JAMA psychiatry 78, 101–109 (2021) (cit. on p. 1).

22. Meisner, A. et al. Combined Utility of 25 Disease and Risk Factor Polygenic Risk Scores for Stratifying Risk of All-Cause Mortality. American Journal of Human Genetics 107, 418–431. issn: 15376605 (2020) (cit. on pp. 1, 6).

23. Jukarainen, S., Kiiskinen, T., Havulinna, A. S. & Karjalainen, J. Genetic risk factors have a substantial impact on healthy life years. medRxiv, 1–55. https://www.medrxiv.org/content/10.1101/2022.01.25.22269831v1 (2022) (cit. on pp. 2, 8).

24. Crawford, D. C., Cooke Bailey, J. N. & Briggs, F. Mind the gap: resources required to receive, process and interpret research-returned whole genome data. Human genetics 138, 691–701 (2019) (cit. on p. 2).

25. Haga, S. B. et al. Public knowledge of and attitudes toward genetics and genetic testing. Genetic testing and molecular biomarkers 17, 327–335 (2013) (cit. on p. 2).

26. Hurle, B. et al. What does it mean to be genomically literate?: National Human Genome Research Institute meeting report. Genetics in Medicine 15, 658–663 (2013) (cit. on p. 2).

27. Lea, D. H., Kaphingst, K. A., Bowen, D., Lipkus, I. & Hadley, D. W. Communicating genetic and genomic information: health literacy and numeracy considerations. Public health genomics 14, 279–289 (2011) (cit. on p. 2).

28. Dwyer, A. A. et al. Evaluating co-created patient-facing materials to increase understanding of genetic test results. Journal of Genetic Counseling 30, 598–605 (2021) (cit. on p. 2).

29. Moscarello, T., Murray, B., Reuter, C. M. & Demo, E. Direct-to-consumer raw genetic data and third-party interpretation services: more burden than bargain? Genetics in Medicine 21, 539–541 (2019) (cit. on p. 2).

30. Davis, K. W., Hamby Erby, L., Fiallos, K., Martin, M. & Wassman, E. R. A comparison of genomic laboratory reports and observations that may enhance their clinical utility for providers and patients. Molecular genetics & genomic medicine 7, e00551 (2019) (cit. on p. 2).

31. Kaye, C. & Korf, B. Genetic literacy and competency. Pediatrics 132, S224–S230 (2013) (cit. on p. 2).

32. Henneman, L., Marteau, T. M. & Timmermans, D. R. Clinical geneticists’ and genetic counselors’ views on the communication of genetic risks: a qualitative study. Patient education and counseling 73, 42–49 (2008) (cit. on p. 2).

33. the alarming rise of complex genetic testing in human embryo selection. Nature 603, 549–550 (2022) (cit. on p. 2).

34. (Cit. on p. 2).

35. Buddeke, J. et al. Comorbidity in patients with cardiovascular disease in primary care: a cohort study with routine healthcare data. eng. Br J Gen Pract 69, e398–e406. issn: 1478-5242 (Electronic); 0960-1643 (Print); 0960-1643 (Linking) (2019) (cit. on p. 2).

36. Of Medicine, I. Cardiovascular Disability: Updating the Social Security Listings isbn: 978-0-309-15698-1. https://nap.nationalacademies.org/catalog/12940/cardiovasculardisability-updating-the-social-security-listings (The National Academies Press, Washington, DC, 2010) (cit. on p. 2).

37. Long, A. N. & Dagogo-Jack, S. Comorbidities of diabetes and hypertension: mechanisms and approach to target organ protection. eng. J Clin Hypertens (Greenwich) 13, 244– 251. issn: 1751-7176 (Electronic); 1524-6175 (Print); 1524-6175 (Linking) (2011) (cit. on p. 2).

38. Bähler, C., Schoepfer, A. M., Vavricka, S. R., Brüngger, B. & Reich, O. Chronic comorbidities associated with inflammatory bowel disease: prevalence and impact on healthcare costs in Switzerland. European Journal of Gastroenterology & Hepatology 29. https://journals.lww.com/eurojgh/Fulltext/2017/08000/Chronic_comorbidities_associated_with_inflammatory.8.aspx 2017) (cit. on p. 2).

39. Wang, J.-H., Wu, Y.-J., Tee, B. L. & Lo, R. Y. Medical Comorbidity in Alzheimer’s Disease: A Nested Case-Control Study. eng. J Alzheimers Dis 63, 773–781. issn: 1875-8908 (Electronic); 1387-2877 (Linking) (2018) (cit. on p. 2).

40. Santiago, J. A. & Potashkin, J. A. The Impact of Disease Comorbidities in Alzheimer’s Disease. eng. Front Aging Neurosci 13, 631770. issn: 1663-4365 (Print); 1663-4365 (Electronic); 1663-4365 (Linking) (2021) (cit. on p. 2).

41. Al-Asadi, A. M., Klein, B. & Meyer, D. Multiple comorbidities of 21 psychological disorders and relationships with psychosocial variables: a study of the online assessment and diagnostic system within a web-based population. Journal of medical Internet research 17, e55–e55. https://pubmed.ncbi.nlm.nih.gov/25803420 (Feb. 2015) (cit. on p. 2).

42. Kessler, R. C., Chiu, W. T., Demler, O., Merikangas, K. R. & Walters, E. E. Prevalence, severity, and comorbidity of 12-month DSM-IV disorders in the National Comorbidity Survey Replication. eng. Arch Gen Psychiatry 62, 617–627. issn: 0003-990X (Print); 1538-3636 (Electronic); 0003-990X (Linking) (2005) (cit. on p. 2).

43. Farabaugh, A. et al. Relationships between major depressive disorder and comorbid anxiety and personality disorders. eng. Compr Psychiatry 46, 266–271. issn: 0010-440X (Print); 0010-440X (Linking) (2005) (cit. on p. 2).

44. Slade, T. & Watson, D. The structure of common DSM-IV and ICD-10 mental disorders in the Australian general population. eng. Psychol Med 36, 1593–1600. issn: 0033-2917 (Print); 0033-2917 (Linking) (2006) (cit. on p. 2).

45. Vollebergh, W. A. et al. The structure and stability of common mental disorders: the NEMESIS study. eng. Arch Gen Psychiatry 58, 597–603. issn: 0003-990X (Print); 0003-990X (Linking) (2001) (cit. on p. 2).

46. Buckley, P. F., Miller, B. J., Lehrer, D. S. & Castle, D. J. Psychiatric comorbidities and schizophrenia. eng. Schizophr Bull 35, 383–402. issn: 0586-7614 (Print); 1745-1701 (Electronic); 0586-7614 (Linking) (2009) (cit. on p. 2).

47. Martin, A. R. et al. Clinical use of current polygenic risk scores may exacerbate health disparities. Nature genetics 51. PMC6563838, 584 (2019) (cit. on p. 3).

48. Privé, F. et al. Portability of 245 polygenic scores when derived from the UK Biobank and applied to 9 ancestry groups from the same cohort. American Journal of Human Genetics 109, 12–23. issn: 15376605 (2022) (cit. on p. 3).

49. Weissbrod, O. et al. Leveraging fine-mapping and multipopulation training data to improve cross-population polygenic risk scores. Nature Genetics 54. issn: 1061-4036 (2022) (cit. on p. 3).

50. Shi, H. et al. Population-specific causal disease effect sizes in functionally important regions impacted by selection. Nature Communications 12. issn: 20411723. http://dx.doi.org/10.1038/s41467-021-21286-1 (2021) (cit. on p. 3).

51. Gugic, J., Zaletel, L. Z. & Oblak, I. Treatment-related Cardiovascular Toxicity in Long-term Survivors of Testicular Cancer. eng. Radiol Oncol 51, 221–227. issn: 1318-2099 (Print); 1581-3207 (Electronic); 1318-2099 (Linking) (2017) (cit. on p. 12).

52. Feldman, D. R. et al. Predicting Cardiovascular Disease Among Testicular Cancer Survivors After Modern Cisplatin-based Chemotherapy: Application of the Framingham Risk Score. Clinical genitourinary cancer 16, e761–e769. https://pubmed.ncbi.nlm.nih.gov/29534941 (Aug. 2018) (cit. on p. 12).

53. Zaid, M. A. et al. Clinical and Genetic Risk Factors for Adverse Metabolic Outcomes in North American Testicular Cancer Survivors. Journal of the National Comprehensive Cancer Network J Natl Compr Canc Netw 16, 257–265 (2018) (cit. on p. 12).

54. Yong, S. Y., Raben, T. G., Lello, L. & Hsu, S. D. Genetic Architecture of Complex Traits and Disease Risk Predictors. Scientific Reports 10. [PMC7374622] (2020) (cit. on p. 14).

