## Supplementary Information for "Polygenic Health Index, General Health, Pleiotropy, Embryo Selection and Disease Risk"

#### 1 Data set

##### 1.1 Phenotype definitions

The disease definitions used various UKB-fields to define case and control status, where an individual with any of the diagnoses or data fields filled was counted as a case for that disease. The definitions used the ICD9, ICD10 and OPCS4 codes in UKB-fields 41271, 41270, 41272; self-reported non-cancer codes from UKB-field 20002 and cancer codes in UKB-field 20001. Additionally, some diseases were specifically included in the intake questionnaire or otherwise used other UKB-fields, which also are listed below.

Most definitions did not use all possible fields such that the UKB was partly underused, i.e., there are cases that incorrectly passed as controls for many of the diseases. There might thus be some quantitative performance gains should the predictors be retrained and validated on more comprehensive phenotype definitions.

Furthermore, training and evaluation of the predictors used different data updates from UKB as more data successively become available. The UKB-trained predictors used a download date of April 2019, whereas the evaluation used data from a download date of April 2021 (AFib and CAD were exceptions using the evaluation data also in training). The latter download date had significantly more cases for some of the diseases and it is these numbers that are reported in the paper. The following disease defintions were used:

**Alzheimer's Disease** ICD10: `F000-F009, G300-G309`

**Asthma** non-cancer codes: `1111`

**Atrial fibrillation** non-cancer codes: `1471, 1483`; ICD10: `I48, I480-I484, I489`; ICD9: `4273`; OPCS4: `K571, K621-K624`

**Basal cell carcinoma** cancer codes: `1061`

**Breast cancer** cancer codes: `1002`

**Coronary artery disease** non-cancer codes: `1075`; ICD9: `410, 4109, 412, 4129`; ICD10: `I21, I210-I214, I219, I21X, I22, I220, I221, I228, I229, I23, I230-I236, I238, I241, I252`; OPCS4: `K401-K404, K411-K414, K451-K455, K491, K492, K498, K499, K502, K751-K754, K758, K759`

**Diabetes type I** ICD10: `E100-E109`

**Diabetes type II** ICD10: `E110-E119`

**Gout** non-cancer codes: `1466`

**Heart attack** non-cancer codes: `1075`

**Hypercholesterolemia** non-cancer codes: `1473`

**Hypertension** non-cancer codes: `1065`

**Inflammatory bowel disease** ICD10: `K500-K509, K510-K519`

**Ischemic stroke** ICD10: `I630-I639`

**Major depressive disorder** ICD10: `F320-F329, F330-339, F340-F349, F380-F389, F390-F399`

**Malignant melanoma** cancer codes: `1059`

**Obesity** UKB-field 2100 where a (weight and height based) BMI over 30 counts a case.

**Prostate cancer** cancer codes: `1044`

**Schizophrenia** ICD10: `F220-F209`; UKB-field: `20544` having coding `2`.

**Testicular cancer** cancer codes: `1045`

#### 1.2 Test set demographics

The test data set consisted of 39,913 self-reported white individuals, 23,110 females and 16,803 males, with mean age 70.4 years at data download (2021) and a standard deviation of 7.3 years. A plot of the age histograms can be found in **Figure 1**.

The disease prevalences in the test set are shown in the same figure, to the right. The UKB population is generally healthier than the general U.S. white population. Furthermore, the disease prevalence in the data set is only an approximation for the lifetime risks, as most participants still may develop any of the conditions in the future. The non-comprehensive

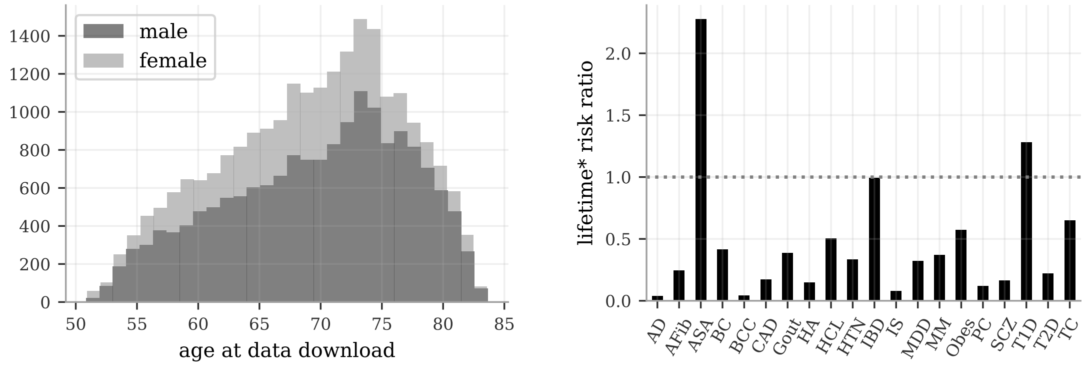

**Figure 1: Age histograms and lifetime* risks of the test set. Left:** The age histograms of the test set for males and females. Most participants are late in life. * Many diseases have late onset however and the UKB lifetime risks are therefore underestimates. Similarly, the evaluations are measuring case status up to data collection without taking age or censoring into account. **Right:** The ratio between UKB lifetime risk estimates and lifetime risks for the white U.S. population, i.e., the lifetime prevalence up to data collection in UKB divided by literature estimates of lifetime risks in the white U.S. population. The ratios are generally much smaller than the reference line at ratio 1.0. The absolute values of both UKB and U.S. prevalences are shown in Figure 2.

disease definitions used also undercount the number of cases in UKB. For the sake of the index construction, we used literature values for the lifetime risks $\rho_d$. Figure 1 shows to the right the UKB prevalences relative to the general white U.S. population. The absolute values for both lifespan impact weights $l_d$ and the lifetime risks $\rho_d$ used in the index can be found in Figure 2.

Note that the frequently used metric RRR is dependent on the prevalence in a selection experiment. This is shown in Figure 3 for theoretical RRR based on a predictor with AUC 0.64. The RRR resulting from a selection experiment decreases with higher prevalence. The precise RRR values are therefore dependent on the absolute prevalences in the population an index is evaluated on.

The index construction also includes the related lifetime risks $\rho_d$ as parameters. We chose literature estimates for the general white U.S. population for these, rather than using the UKB prevalences as estimates.

### 2 Predictor Specifics

#### 2.1 Individual Predictor Construction

Most of the predictors used in this paper were trained with the LASSO algorithm on the UK Biobank, using the methods described in Lello et al. [67, 68]. Several other disease conditions were trained using the PRS-CS package [69, 70] and the EUR 1000 Genomes reference panel coupled with a publicly available GWAS. For these traits, GWAS were selected that specifically excluded the UK Biobank participants in the GWAS to prevent inflated performance metrics. The GWAS were pruned by filtering down to markers which are present in the UK Biobank calls before running PRS-CS with the 1000 Genomes EUR LD panels. In addition to LASSO

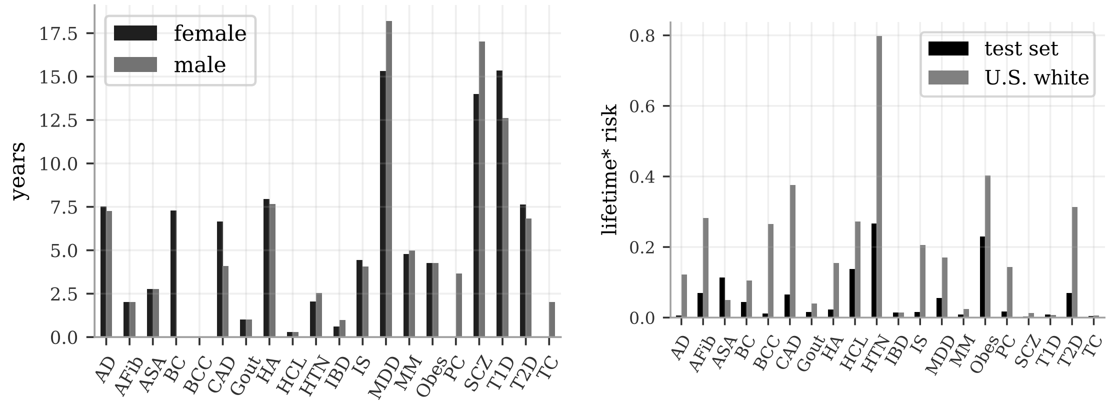

**Figure 2: The weights $l_d$, average number of life years lost due to the disease, and the lifetime$^*$ risks $\rho_d$ used to construct the index. Left:** The weights $l_d$ in the main text equation (1) are estimates of life years lost due to having a disease as compared to the general population lifespan, as deduced from literature studies [1–66]. **Right:** The lifetime disease risks for the UKB data compared to the U.S. general white population. The UKB numbers are the lifetime prevalence up to data collection are hence underestimates. The risks were averaged over the sexes except for BC, PC and TC. We used the values for the white U.S. population in the index construction in main text equation (1). The ratio between UKB and U.S. risks are also shown to the right in **Figure 1**

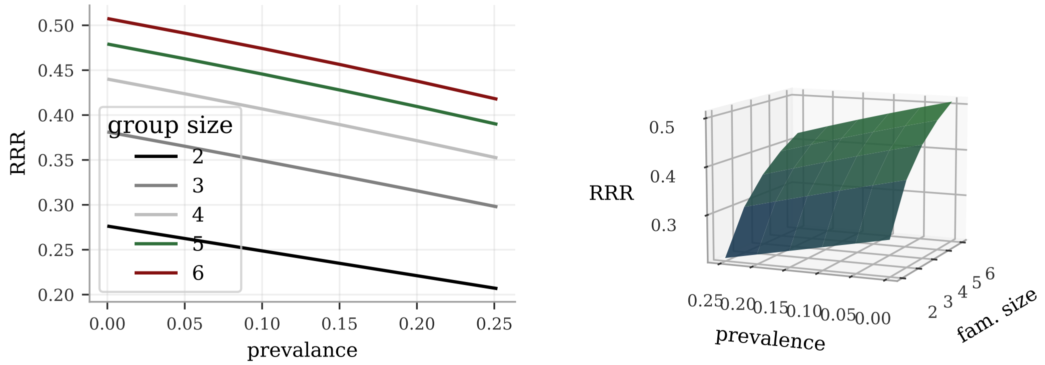

**Figure 3: The RRR for selection experiments depend on the prevalence in the population.** The RRR for selecting on a single predictor can be calculated theoretically using the Gaussian risk model. The metric varies with disease prevalence with lower RRR for more common diseases. This example used the fairly typical AUC of .64.

and PRS-CS, we used a publicly available schizophrenia predictor which was then filtered to markers which overlap the UK Biobank imputed set and filtered for p-value $< 0.05$ resulting in 24,387 markers. We now list the construction methods and data sources for each predictor along with the AUC on the testing set described in 1.2.

**Alzheimer's Disease** GWAS [71] + PRS-CS [69, 70] - 21,982 European ancestry cases, 41,944 European ancestry controls; retreived from [72]. AUC: $.686 \pm .004$

**Asthma** UK Biobank LASSO [67, 68] - trained on 48,875 cases, 369,158 controls. Hyperparameter selection on 500 cases and 500 controls. AUC: $.626 \pm .004$

**Atrial fibrillation** UK Biobank LASSO [67, 68] - trained on 29,206 cases, 388,670 controls. Hyperparameter selection on 500 cases and 500 controls. AUC: $.623 \pm .004$

**Basal cell carcinoma** UK Biobank LASSO [67, 68] - trained on 3,795 cases, 414,238 controls. Hyperparameter selection on 500 cases and 500 controls. AUC: $.618 \pm .011$

**Breast cancer** UK Biobank LASSO [67, 68] - trained on 9,459 cases, 216,339 controls. Hyperparameter selection on 100 cases and 100 controls. AUC: $.594 \pm .008$

**Coronary artery disease** UK Biobank LASSO [67, 68] - trained on 27,172 cases, 390,704 controls. Hyperparameter selection on 500 cases and 500 controls. AUC: $.616 \pm .005$

**Diabetes type I** UK Biobank LASSO [67, 68] - trained on 2,345 cases, 415,688 controls. Hyperparameter selection on 500 cases and 500 controls. AUC: $.627 \pm .015$

**Diabetes type II** UK Biobank LASSO [67, 68] - trained on 18,097 cases, 399,936 controls. Hyperparameter selection on 500 cases and 500 controls. AUC: $.616 \pm .004$

**Gout** UK Biobank LASSO [67, 68] - trained on 5,712 cases, 412,321 controls. Hyperparameter selection on 500 cases and 500 controls. AUC: $.654 \pm .011$

**Heart attack** UK Biobank LASSO [67, 68] - trained on 9,455 cases, 408,578 controls. Hyperparameter selection on 500 cases and 500 controls. AUC: $.580 \pm .008$

**Hypercholesterolemia** UK Biobank LASSO [67, 68] - trained on 53,603 cases, 364,430 controls. Hyperparameter selection on 500 cases and 500 controls. AUC: $.616 \pm .003$

**Hypertension** UK Biobank LASSO [67, 68] - trained on 110,893 cases, 307,140 controls. Hyperparameter selection on 500 cases and 500 controls. AUC: $.635 \pm .003$

**Inflammatory bowel disease** GWAS [73] + PRS-CS [69, 70] - 12k cases + 21k controls - retrieved from [74]. AUC: $.647 \pm .003$

**Ischemic stroke** GWAS [75] + PRS-CS [69, 70] - 67,162 cases and 454,450 controls, retrieved from [76]. AUC: $.541 \pm .002$

**Major depressive disorder** GWAS [77] + PRS-CS [69, 70] - 45,396 cases and 97,250 controls - retrieved `daner_PGC_MDD_noUKB_no23andMe.txt` from [78]. AUC: $.534 \pm .001$

**Malignant melanoma** UK Biobank LASSO [67, 68] - trained on 2,911 cases, 415,122 controls. Hyperparameter selection on 500 cases and 500 controls. AUC: $.573 \pm .016$

**Obesity** UK Biobank LASSO [67, 68] - trained on 417,687 continuous measurements from field 2100 with 1,000 used for hyperparameter selection. Predictor then evaluated on the test set using the Obesity definition. AUC: .669 ± .002

**Prostate cancer** UK Biobank LASSO [67, 68] - trained on 3,275 cases, 189,560 controls. Hyperparameter selection on 100 cases and 100 controls. AUC: .636 ± .015

**Schizophrenia** The schizophrenia predictor was obtained using the results from [79]. Retrieved the file `scz2.prs.txt.gz` from [80]. The predictor was filtered to SNPs which overlap the UKB imputed set and filtered for p-value $< 0.05$ resulting in 24,387 SNPs. The beta value was calculated as the log(OR). AUC: .673 ± .029

**Testicular cancer** UK Biobank LASSO [67, 68] - trained on 650 cases, 192,185 controls. Hyperparameter selection on 100 cases and 100 controls. AUC: .612 ± .041

##### 2.2 AUC evaluation

The uncertainties in AUC for each predictor in **Table 1** in the main text were computed via the following algorithm. Case/control numbers and mean PRS were computed in the test set and a theoretical PRS-distribution was defined, according to equation (2) in the main text. The same numbers of cases and controls that were in the test set were sampled from the case and control parts of the PRS-distribution, respectively. An AUC was computed based on the sampled PRS and the procedure was repeated 30 times. The standard deviation from these repeated computations is the error reported next to the AUC.

#### 3 Additional Selection Experiments

##### 3.1 Selection experiments in genetic trios

The full RRR and index gain plots for the index selection among genetic trios are shown in **Figure 4**. Note that the error bars are very large and most disease RRR and index gains are inconclusive in this experiment. We also display a comparison of total index gain in DALY for pairs and trios of both siblings and unrelated individuals in **Figure 5**.

##### 3.2 Sex bias adjusted health index

The index is defined with sex specific parameters $l_d$ and $\rho_d$ and includes different diseases for males (PC, TC) and females (BC). Consequently, the health index distributions are somewhat different for the two sexes. The effect is small but existant, as can be seen in **Figure 6**. The selection experiments are sensitive to this and the larger the group size the stronger is the dependence on the right tails, i.e., on the distribution differences for the highest health index values. As can be seen to the left in the figure, there is a larger proportion of females than males in the test set with very high health index as compared to the intermediate or lower index value regions. This is a result of the particular choice of index and test set but comports well in both direction and scale of general life expectancy differences. As a result, however, direct selection on the health index leads to an over-representation of women in the selected set. We defined a minimal non-linear transformation of the male and female health index

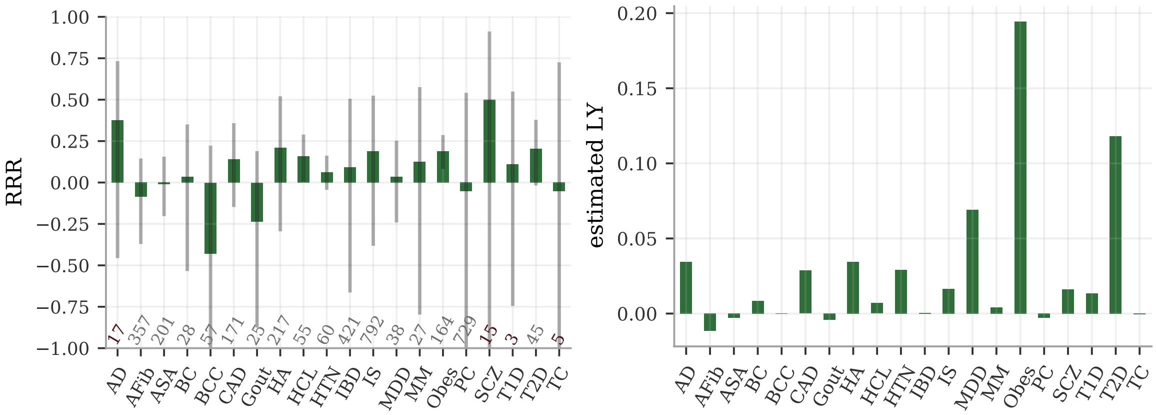

**Figure 4: Index selection between 969 trios of genetic siblings. Left:** The RRR result is inconclusive for most diseases, as is seen by the theoretical error bars (using 95% C.I. from Wilson score interval applied to the selected prevalences); the figure is cropped at RRR $= \pm 1$. The small sample size of trios is enough only to statistically determine non-zero RRR for HCL and Obes, while HTN and T2D borders to significance. All these are positive. **Right:** The index gain from the selection among the trios shows no strong negative components. No error bars were computed but the uncertainties are naturally very large for the trios also in this metric.

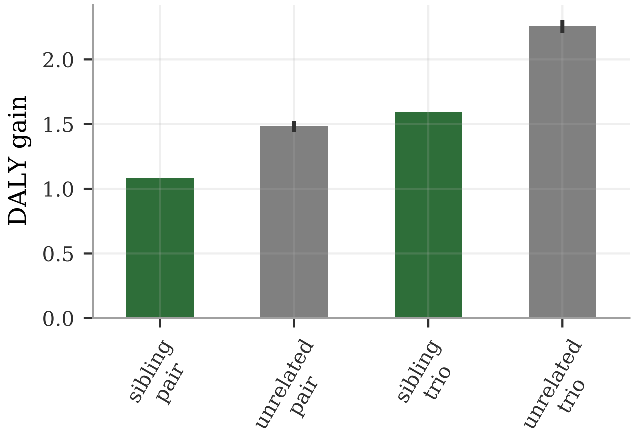

**Figure 5: Index gain in DALY from selection among pairs and trios of siblings and unrelated individuals.** Selection among siblings retains most of the gain for both sibling pairs and trios, as compared to selection among unrelated individuals. The sibling/unrelated ratios are .73 and .71 for pairs and trios, respectively. The error bars for the unrelated individuals are 95% C.I. estimates from 25 selection experiments. No error bars were computed for the sibling results but the uncertainties are larger than for the selection among unrelated individuals.

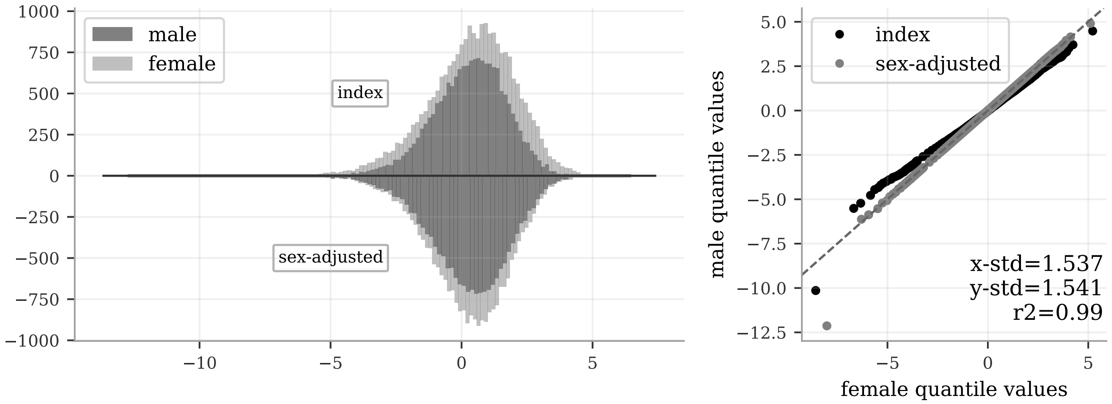

**Figure 6: The health index histograms for males and females, before and after sex-adjustment. Left:** The health index histograms for females and males are plotted on the positive y-axis, while the sex-adjusted histograms are plotted on the negative axis. There were more females (23,110) in the test set than males (16,803). The adjustment is minor but with noticeable effect on the tails; the corresponding densities (normalizing by total number of females/males) are practically identical after the sex adjustment. **Right:** A QQ-plot of the female and male health index distributions before and after sex-adjustment. The plotted dots correspond to percentiles but with extra focus on the tails; the 0-3 percentiles and 97-100 percentiles are split into 40 equidistant points each such that the tail behaviors are shown clearly. The sex-adjusted distributions agree almost exactly, with a regression $R^2$ of 0.99 (affected only by the extreme outlier at 0.075th percentile). As such, a sex adjusted health index could therefore be used to compare the health of males to females without preference to either, as both are measured relative to their respective cohorts.

values mapping them to their mean distribution for a sex neutral health index. The result is plotted on the negative y-axis in Figure 6, with the resulting QQ-plot to the right. Selecting on the sex-adjusted index kept the females-to-males ratio among the selected equal to the total test set ratio. This had minor measurable effects on the index performance and the results for group size five are shown in Figure 7.

#### 3.3 Non-European ancestry

The main part of this paper dealt exclusively with a data set of European ancestry. All predictors were trained on such a cohort and it is a well-established fact that predictor performance declines with the genetic distance between two populations (typically linearly when measured in $R^2$, see for example [81]). Nevertheless, some of the performance of the Euro-trained predictors is retained when applied to other ancestries and we demonstrate here that even a composite health index has non-trivial performance for people of South Asian (SAS), East Asian (EAS), and African (AFR) ancestry. Based on self-reported ancestry in UKB, we created test sets with 9,438 (SAS), 1,493 (EAS), and 7,614 (AFR) samples, withheld from all training and hyperparameter tuning.

We used the same type of index construction but excluded basal cell carcinoma and malignant melanoma because these are close to non-existant diseases in these test sets and major depressive disorder because its poor individual predictor performance.

For each test set, we used ancestry specific weights and population risks $l_d, \rho_d$. The individual disease RRR and index component gains for SAS are shown in Figure 8 for selection

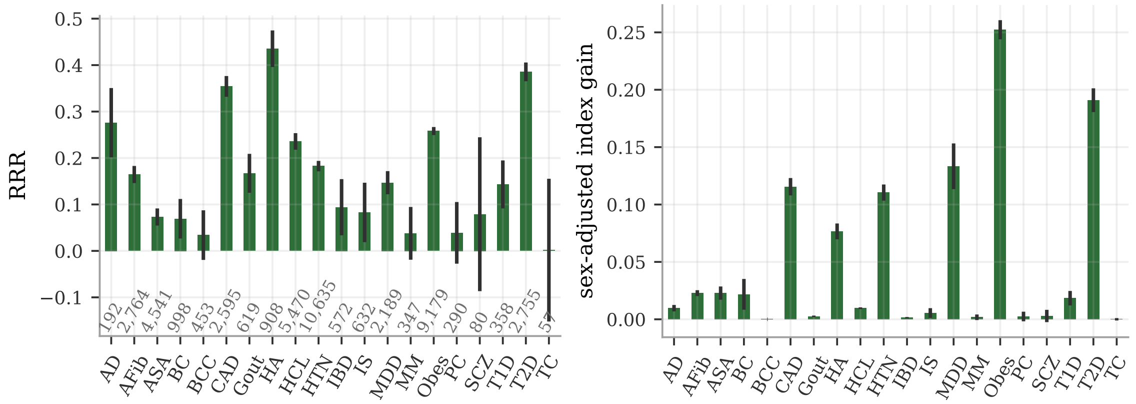

**Figure 7: The RRR and index gain for selection on the sex-adjusted health index for group size of five. Left:** The RRR values are generally similar when selecting on the sex-adjusted health index. As in the main document, the case numbers are listed just above the x-axis. The error bars are 95% C.I. estimates from 25 selection experiments. **Right:** The index gain is also just slightly affected with no qualitative differences. Since the sex-adjustment is a non-linear transformation, it is no longer technically possible to interpret the gain in life years. The error bars are again bootstrap estimates of the 95% C.I. from 25 selection experiments.

among groups of size 5, while the total index gains are shown for EAS and AFR in Figure 9. The RRR result is overwhelmingly positive also for South Asian ancestry, again reaching over or about 40% for a couple of traits (AD, HA). Notably, the Alzheimer's disease risk was reduced more for SAS than EUR in these experiments even taking the large error bars into account. This is based on only 34 SAS AD cases, however. The case numbers are always included above the x-axis in the plots for this reason. Another observation is the differences in type II diabetes. Although still with a strong relative risk reduction of 18%, the SAS result is about half the RRR of the EUR index. The SAS RRR for IBD is also worse and appears to have a borderline statistically significant negative mean value. As seen in Figure 10 below, the IBD predictor trained in EUR has more negative pairwise correlations with other disease PRS when applied to SAS. In particular BC, SCZ, T1D and T2D, with their much stronger index weights, may counter the predicted IBD risk for SAS. To be clear, the PRS correlations in the SAS data sets still refer to the predictors trained in the EUR data set and the differences may be due to distinct linkage disequilibrium patterns in general; it is still unknown what the PRS correlations would be using a training set of SAS ancestry. Lastly, we note that MDD still has a significant positive RRR despite the fact that there was no direct MDD PRS included in the South Asian health index.

The SAS index gains for the components are overwhelmingly positive and dominated by CAD, heart attack, hypertension, major depressive disorder, obesity, and type II diabetes. Again, we note that MDD is contributing a lot — due to its high prevalence and strong impact — despite not being in the index directly. There was no statistically significant negative contributions to the SAS index.

The index gain from selection among EAS and AFR also performs well, as seen in Figure 9. We detect a measurable attenuation from the EUR result, as is expected due to the genetic distance from the European training population. Yet, there is a consistent and strongly significant positive gain for both EAS and AFR when using the EUR-trained predictors and

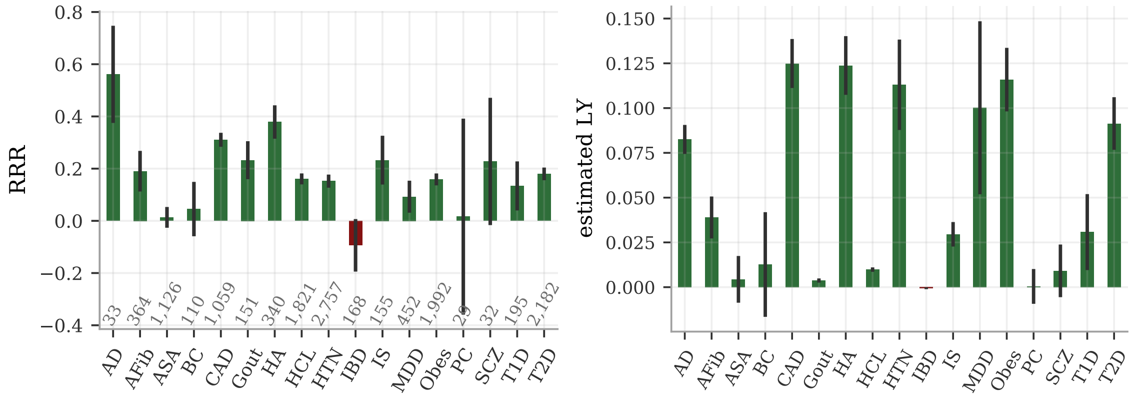

**Figure 8: The RRR and index gain for selection on a South Asian health index among unrelated groups of size five.** In both figures, the error bars are 95% C.I. estimates from 25 selection experiments. **Left:** The RRR from the index selection is overwhelmingly positive also in the SAS test set. The borderline statistically significant negative RRR for IBD is the most notable difference from the EUR result and can be traced to IBD's more negative PRS-correlations with other predictors in SAS as compared to EUR (see Figure 10). Note again that the PRS is computed by the EUR trained predictor applied to SAS and may reflect population differences in linkage disequilibrium rather than underlying biology in SAS. **Right:** The component-wise index gain is also predominantly positive. The possibly negative RRR for IBD has almost no impact at all on the index due to its small weight and low prevalence.

ancestry specific parameters in the index construction.

The phenotypic and genetic correlation characterization of the diseases and predictors in the South Asian test set is shown in Figure 10.

### References

1. Albertson, P. C. Long-term Survival Among Men With Conservatively Treated Localized Prostate Cancer. *JAMA: The Journal of the American Medical Association* **274,** 626 (Aug. 1995) (cit. on p. 4).
2. Alcocer, L. & Cueto, L. Hypertension, a health economics perspective. *Therapeutic Advances in Cardiovascular Disease* **2,** 147–155 (June 2008) (cit. on p. 4).
3. Berry, J. D. *et al.* Lifetime risks of cardiovascular disease. *New England Journal of Medicine* **366,** 321–329 (Jan. 2012) (cit. on p. 4).
4. Botta, L. *et al.* Changes in life expectancy for cancer patients over time since diagnosis. *Journal of Advanced Research* **20,** 153–159 (Nov. 2019) (cit. on p. 4).
5. Bucholz, E. M. *et al.* Life expectancy and years of potential life lost after acute myocardial infarction by sex and race: a cohort-based study of Medicare beneficiaries. *Journal of the American College of Cardiology* **66,** 645–655 (Aug. 2015) (cit. on p. 4).
6. Bucholz, E. M., Ma, S., Normand, S.-L. T. & Krumholz, H. M. Race, Socioeconomic Status, and Life Expectancy After Acute Myocardial Infarction. *Circulation* **132,** 1338=1346 (Oct. 2015) (cit. on p. 4).

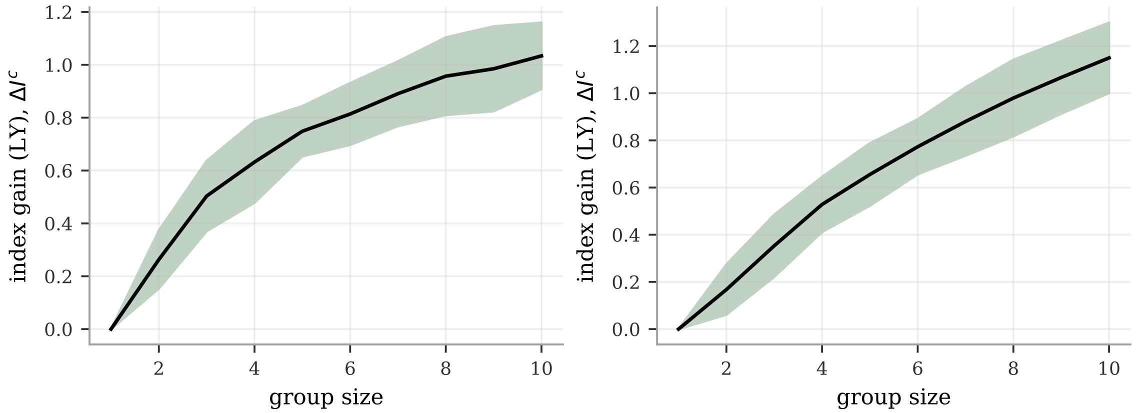

**Figure 9: Index gain from selection experiments as function of group size when the index are applied to East Asian and African test sets.** The selection was done on indices using EUR-trained predictors (excluding BCC, MM, and MDD) but ancestry specific lifetime risks $\rho_d$ and lifespan impact weights $l_d$. Error bars are 95% C.I. as estimated from 25 separate experiments for each ancestry. **Left:** EAS using a test set of 1,493 samples. **Right:** AFR using a test set of 7,614 samples.

7. Burisch, J., Jess, T., Martinato, M. & Lakatos, P. The burden of inflammatory bowel disease in Europe. *Journal of Crohn's and Colitis* **7** (June 2013) (cit. on p. 4).
8. Carson, A. P. *et al.* Ethnic differences in hypertension incidence among middle-aged and older adults: the multi-ethnic study of atherosclerosis. *Hypertension* **57,** 1101–1107 (June 2011) (cit. on p. 4).
9. Carstensen, B., Rønn, P. F. & Jørgensen, M. E. Lifetime risk and years lost to type 1 and type 2 diabetes in Denmark, 1996–2016. *BMJ Open Diabetes Research & Care* **9** (June 2021) (cit. on p. 4).
10. Chang, C.-K. *et al.* Life Expectancy at Birth for People with Serious Mental Illness and Other Major Disorders from a Secondary Mental Health Care Case Register in London. *PLoS ONE* **6** (May 2011) (cit. on p. 4).
11. Chen, V. *et al.* Lifetime Risks for Hypertension by Contemporary Guidelines in African American and White Men and Women. *JAMA Cardiology* **4,** 455 (Mar. 2019) (cit. on p. 4).
12. Coleman, M. P. Opinion: why the variation in breast cancer survival in Europe? *Breast Cancer Research* **1** (Oct. 1999) (cit. on p. 4).
13. Concannon, P. *et al.* Type 1 Diabetes: Evidence for Susceptibility Loci from Four Genome-Wide Linkage Scans in 1, 435 Multiplex Families. *Diabetes* **54,** 2995–3001 (Oct. 2005) (cit. on p. 4).
14. Crump, C., Winkleby, M. A., Sundquist, K. & Sundquist, J. Comorbidities and Mortality in Persons With Schizophrenia: A Swedish National Cohort Study. *American Journal of Psychiatry* **170,** 324–333 (Mar. 2013) (cit. on p. 4).
15. Flohil, S. C. *et al.* Trends in Basal cell carcinoma incidence rates: a 37-year Dutch observational study. *Journal of Investigative Dermatology* **133,** 913–918 (Apr. 2013) (cit. on p. 4).

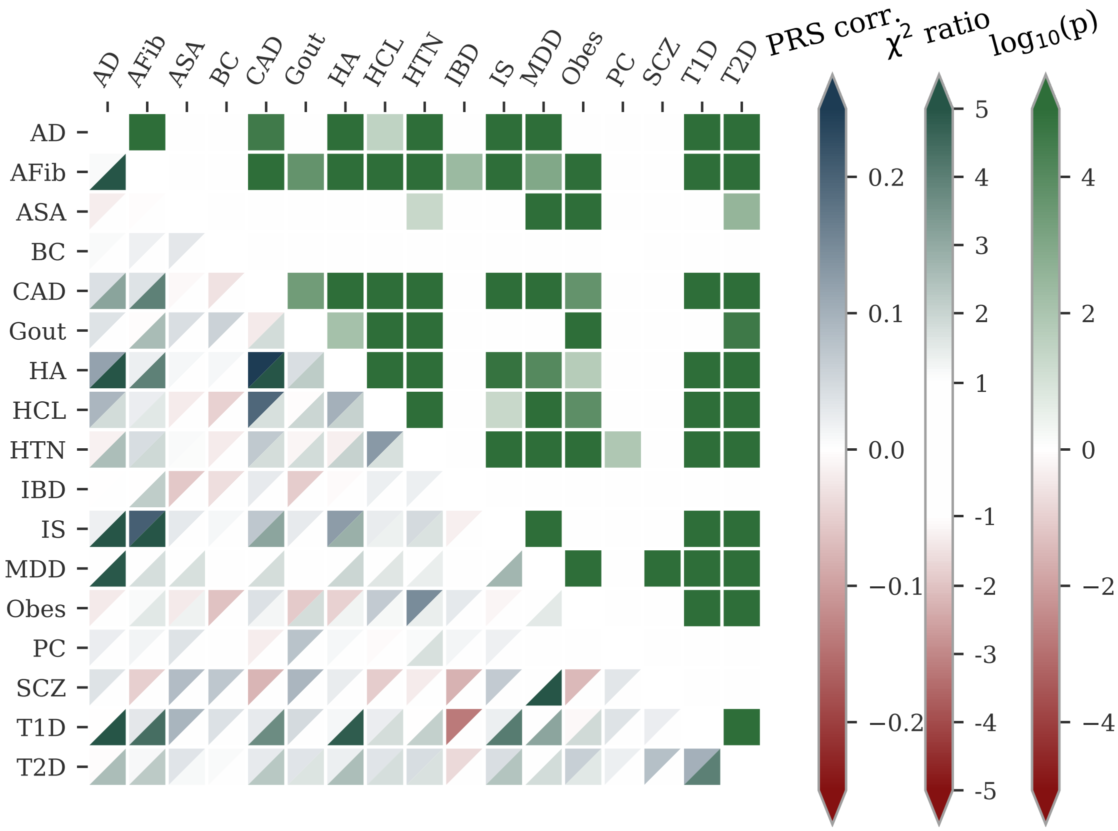

**Figure 10: PRS correlations and phenotypic comorbities in the South Asian test set.** See the main text and **Figure 8** in the paper on how to interpret this figure. The qualitative observations for the EUR ancestry are true for the SAS test set too: the studied diseases tend overwhelmingly to have positive comorbidity with one or more of the other diseases and the PRS are mostly uncorrelated or mildly positive correlated. There are a few more weakly anti-correlated pairs for SAS than for EUR, in particular for schizophrenia and IBD. The latter provides an explanation to why IBD has a worse RRR for SAS than for EUR. This may be an artifact of using EUR trained predictors on SAS ancestry and does not need to reflect the underlying genetic effects and biology. An index built from SAS trained predictor could and will answer such questions as soon as sufficient data is available.

16. Franco, O. H., Peeters, A., Bonneux, L. & de Laet, C. Blood Pressure in Adulthood and Life Expectancy With Cardiovascular Disease in Men and Women. *Hypertension* **46,** 280–286 (June 2005) (cit. on p. 4).

17. Gitsels, L. A., Kulinskaya, E. & Steel, N. Survival prospects after acute myocardial infarction in the UK: a matched cohort study 1987-2011. *BMJ Open* **7** (Dec. 2017) (cit. on p. 4).

18. Gregg, E. W. *et al.* Trends in lifetime risk and years of life lost due to diabetes in the USA, 1985–2011: A modelling study. *The Lancet Diabetes & Endocrinology* **2,** 867–874 (2014) (cit. on p. 4).

19. Hjorthøj, C., Stürup, A. E., McGrath, J. J. & Nordentoft, M. years of potential life lost and life expectancy in schizophrenia: a systematic review and meta-analysis. *The Lancet Psychiatry* **4,** 295–301 (Feb. 2017) (cit. on p. 4).

20. Hogg, R. S., Schechter, M. T., Montaner, J. S. & Hogg, J. C. Asthma mortality in Canada, 1946 to 1990. *Canadian Respiratory Journal* **2,** 61–66 (1995) (cit. on p. 4).

21. Hruby, A. & Hu, F. B. The Epidemiology of Obesity: A Big Picture. *PharmacoEconomics* **33,** 673–689 (July 2014) (cit. on p. 4).

22. Hung, M.-C., Ekwueme, D. U., Rim, S. H. & White, A. Racial/ethnicity disparities in invasive breast cancer among younger and older women: An analysis using multiple measures of population health. *Cancer Epidemiology* **45,** 112–118 (Oct. 2016) (cit. on p. 4).

23. Hutchinson, G *et al.* Morbid Risk of Schizophrenia in First-Degree Relatives of White and African-Caribbean Patients with Psychosis. *British Journal of Psychiatry* **169,** 776–780 (Jan. 1996) (cit. on p. 4).

24. Karjalainen, S., Salo, H. & Teppo, L. Basal Cell and Squamous Cell Carcinoma of the Skin in Finland. *International Journal of Dermatology* **28,** 445–450 (Sept. 1989) (cit. on p. 4).

25. Kassaï, B., Boissel, J.-P., Cucherat, M., Boutitie, F. & Gueyffier, F. Treatment of High Blood Pressure and Gain in Event-Free Life Expectancy. *Vascular Health and Risk Management* **1,** 163–169 (June 2005) (cit. on p. 4).

26. Katzmarzyk, P. T. & Staiano, A. E. New race and ethnicity standards: Elucidating health disparities in diabetes. *BMC Medicine* **10** (Apr. 2012) (cit. on p. 4).

27. Kendler, K. S. The Roscommon Family Study: I. Methods, Diagnosis of Probands, and Risk of Schizophrenia in Relatives. *Archives of General Psychiatry* **50,** 527 (July 1993) (cit. on p. 4).

28. Kharazmi, E. *et al.* Cancer Risk in Relatives of Testicular Cancer Patients by Histology Type and Age at Diagnosis: A Joint Study from Five Nordic Countries. *European Urology* **68,** 283–289 (Aug. 2015) (cit. on p. 4).

29. Kirchgesner, J. *et al.* Impact on Life Expectancy of Withdrawing Thiopurines in Patients with Crohn's Disease in Sustained Clinical Remission: A Lifetime Risk-Benefit Analysis. *PLOS ONE* **11** (June 2016) (cit. on p. 4).

30. Kolomisky-Rabas, P. L. *et al.* Lifetime Cost of Ischemic Stroke in Germany: Results and National Projections From a Population-Based Stroke Registry. *Stroke* **37,** 1179–1183 (Mar. 2006) (cit. on p. 4).

31. Kuenzig, E. M., Manuel, D. G., Donelle, J. & Benchimol, E. I. Life expectancy and health-adjusted life expectancy in people with inflammatory bowel disease. *Canadian Medical Association Journal* **192** (2020), month=11) (cit. on p. 4).

32. Kurian, A. W., Fish, K., Shema, S. J. & Clarke, C. A. Lifetime risks of specific breast cancer subtypes among women in four racial/ethnic groups. *Breast Cancer Research* **12** (Nov. 2010) (cit. on p. 4).

33. Lange, P., Çolak, Y., Ingebrigtsen, T. S., Vestbo, J. & Marott, J. L. Long-term prognosis of asthma, chronic obtrusive pulmonary disease, and asthma-chronic obstructive pulmonary disease overlap in the Copenhagen City Heart study: a prospective population-based analysis. *The Lancet Respiratory Medicine* **4,** 454–462 (Apr. 2016) (cit. on p. 4).

34. Larson, E. B. *et al.* Survival after Initial Diagnosis of Alzheimer Disease. *Annals of Internal Medicine* **140,** 501 (Apr. 2004) (cit. on p. 4).

35. Laursen, T. M. Life expectancy among persons with schizophrenia or bipolar affective disorder. *Schizophrenia Research* **131,** 101–104 (Sept. 2011) (cit. on p. 4).

36. Laursen, T. M., Musliner, K. L., Benros, M. E., Vestergaard, M. & Munk-Olsen, T. Mortality and life expectancy in persons with severe unipolar depression. *Journal of Affective Disorders* **193,** 203–207 (Mar. 2016) (cit. on p. 4).

37. Leening, M. J. *et al.* Sex differences in lifetime risk and first manifestation of cardiovascular disease: prospective population based cohort study. *BMJ* **349** (Nov. 2014) (cit. on p. 4).

38. Livingstone, S. J. *et al.* Estimated Life Expectancy in a Scottish Cohort With Type 1 Diabetes, 2008-2010. *JAMA* **313,** 37 (Jan. 2015) (cit. on p. 4).

39. Lloyd-Jones, D. M., Larson, M. G., Beiser, A. & Levy, D. Lifetime risk of developing coronary heart disease. *The Lancet* **353,** 89–92 (Jan. 1999) (cit. on p. 4).

40. Lloyd-Jones, D. M. *et al.* Lifetime Risk for Developing Congestive Heart Failure: The Framingham Heart Study. *Circulation* **106,** 3068–3072 (Dec. 2002) (cit. on p. 4).

41. LLoyd-Jones, D. M. *et al.* Prediction of Lifetime Risk for Cardiovascular Disease by Risk Factor Burden at 50 Years of Age. *Circulation* **113,** 791–798 (Feb. 2006) (cit. on p. 4).

42. Lloyd, T. *et al.* Lifetime risk of being diagnosed with, or dying from, prostate cancer by major ethnic group in England 2008-2010. *BMC Medicine* **13** (July 2015) (cit. on p. 4).

43. Lobo, A *et al.* Incidence and lifetime risk of dementia and Alzheimer's disease in a Southern European population. *Acta Psychiatrica Scandinavica* **124,** 372–383 (Aug. 2011) (cit. on p. 4).

44. Mirabelli, M. C., Beavers, S. F., Chatterjee, A. B. & Moorman, J. E. Age at asthma onset and subsequent asthma outcomes among adults with active asthma. *Respiratory Medicine* **107,** 1829–1836 (Dec. 2013) (cit. on p. 4).

45. Morgan, C. L., Currie, C. J. & Peters, J. R. Relationship between diabetes and mortality: a population study using record linkage. *Diabetes Care* **23,** 1103–1107 (Aug. 2000) (cit. on p. 4).

46. Mou, L. *et al.* Lifetime Risk of Atrial Fibrillation by Race and Socioeconomic Status. *Circulation: Arrhythmia and Electrophysiology* **11** (July 2018) (cit. on p. 4).

47. Narayan, K. M., Boyle, J. P., Thompson, T. J., Sorensen, S. W. & Williamson, D. F. Lifetime risk for diabetes mellitus in the United States. *JAMA* **290,** 1884 (Oct. 2003) (cit. on p. 4).

48. Pedersen, C. B. *et al.* A Comprehensive Nationwide Study of the Incidence Rate and Lifetime Risk for Treated Mental Disorder. *JAMA Psychiatry* **71,** 573 (May 2014) (cit. on p. 4).

49. Collaboration, P. S. Body-mass index and cause-specific mortality in 900 000 adults: collaborative analyses of 57 prospective studies. *The Lancet* **373,** 1083–1096 (Mar. 2009) (cit. on p. 4).

50. Rawshani, A. *et al.* Excess mortality and cardiovascular disease in type 1 diabetes in relation to age at onset: a nationwide study of 27, 195 young adults with diabetes. *The Lancet* **392,** 477–486 (Aug. 2018) (cit. on p. 4).

51. Redondo, M. J. Genetics of type 1A diabetes. *Recent Progress in Hormone Research* **56,** 69–90 (Jan. 2001) (cit. on p. 4).

52. Salama, A. K. *et al.* The Effect of Metastatic Site and Decade of Diagnosis on the Individual Burden of Metastatic Melanoma: Contemporary Estimates of Average Years of Life Lost. *Cancer Investigation* **30,** 637–641 (Sept. 2012) (cit. on p. 4).

53. Seshadri, S. & Wolf, P. A. Lifetime risk of stroke and dementia: current concepts, and estimates from the Framingham Study. *The Lancet Neurology* **6,** 1106–1114 (Dec. 2007) (cit. on p. 4).

54. Smith, A. J., Lambert, P. C. & Rutherford, M. J. Understanding the impact of sex and stage differences on melanoma cancer patient survival: a SEER-based study. *British Journal of Cancer* **124,** 671–677 (Nov. 2020) (cit. on p. 4).

55. Staerk, L. *et al.* Lifetime risk of atrial fibrillation according to optimal, borderline, or elevated levels of risk factors: cohort study based on longitudinal data from the Framingham Heart Study. *BMJ* **361** (Apr. 2018) (cit. on p. 4).

56. Steensma, C *et al.* Describing the population health burden of depression: health-adjusted life expectancy by depression status in Canada. *Health Promotion and Chronic Disease Prevention in Canada* **36,** 205–213 (Oct. 2016) (cit. on p. 4).

57. Sullivan, P. F., Neale, M. C. & Kendler, K. S. Genetic epidemiology of major depression: review and meta-analysis. *American Journal of Psychiatry* **157,** 1552–1562 (Oct. 2000) (cit. on p. 4).

58. Syriopoulou, E., Bower, H., Andersson, T. M.-L., Lambert, P. C. & Rutherford, M. J. Estimating the impact of a cancer diagnosis on life expectancy by socio-economic group for a range of cancer types in England. *British Journal of Cancer* **117,** 1419–1426 (Oct. 2017) (cit. on p. 4).

59. Thiam, A., Zhao, Z., Quinn, C. & Barber, B. Years of life lost due to metastatic melanoma in 12 countries. *Journal of Medical Economics* **19,** 259–264 (Oct. 2015) (cit. on p. 4).

60. To, T., Wang, C., Guan, J., McLimont, S. & Gershon, A. S. What Is the Lifetime Risk of Physician-diagnosed Asthma in Ontario, Canada? *American Journal of Respiratory and Critical Care Medicine* **181,** 337–343 (2010) (cit. on p. 4).

61. Tsevat, J, Weinstein, M. C., Williams, L. W., Tosteson, A. N. & Goldman, L. Expected gains in life expectancy from various coronary heart disease risk factor modifications. *Circulation* **83,** 1194–1201 (Dec. 1991) (cit. on p. 4).

62. Vasan, R. S., Pencina, M. J., Cobain, M., Freiberg, M. S. & D'Agostino, R. B. Estimated Risks for Developing Obesity in the Framingham Heart Study. *Annals of Internal Medicine* **143,** 473 (Oct. 2005) (cit. on p. 4).

63. Vinter, N. *et al.* Trends in excess mortality associated with atrial fibrillation over 45 years (Framingham Heart Study): community based cohort study. *BMJ* **370,** m2724 (Aug. 2020) (cit. on p. 4).

64. Wong, C. S., Strange, R. C. & T, L. J. Basal cell carcinoma. *BMJ* **327,** 794–798 (Oct. 2003) (cit. on p. 4).

65. Wright, A. K. *et al.* Life Expectancy and Cause-Specific Mortality in Type 2 Diabetes: A Population-Based Cohort Study Quantifying Relationships in Ethnic Subgroups. *Diabetes Care* **40,** 338–345 (Dec. 2016) (cit. on p. 4).

66. Zanetti, O, Solerte, S. B. & Cantoni, F. Life expectancy in alzheimer's disease (AD). *Archives of Gerontology and Geriatrics* **49,** 237–243 (2009) (cit. on p. 4).

67. Lello, L. *et al.* Accurate genomic prediction of human height. *Genetics* **210.** [PMC6216598], 477–497 (2018) (cit. on pp. 3, 5, 6).

68. Lello, L., Raben, T. G., Yong, S. Y., Tellier, L. C. & Hsu, S. D. H. Genomic prediction of 16 complex disease risks including heart attack, diabetes, breast and prostate cancer. *Sci Rep* **9.** [PMC6814833], 1–16 (2019) (cit. on pp. 3, 5, 6).

69. Ge, T., Chen, C.-Y., Ni, Y., Feng, Y.-C. A. & Smoller, J. W. Polygenic prediction via Bayesian regression and continuous shrinkage priors. *Nature communications* **10,** 1–10 (2019) (cit. on pp. 3, 5).

70. *PRScs GitHub repository* `https://github.com/getian107/PRScs`. Accessed: 2022-Feb-22 (cit. on pp. 3, 5).

71. Kunkle, B., Grenier-Boley, B. & Sims, R. e. a. Genetic meta-analysis of diagnosed Alzheimer's disease identifies new risk loci and implicates A$\beta$, tau, immunity and lipid processing. *Nature Genetics* **51,** 414–430. `https://www.nature.com/articles/s41588-019-0358-2` (2019) (cit. on p. 5).

72. *Downloadable Alzheimer's predictor* `http://ftp.ebi.ac.uk/pub/databases/gwas/summary_statistics/GCST007001-GCST008000/GCST007511/Kunkle_etal_Stage1_results.txt`. Accessed: 2022-05-30 (cit. on p. 5).

73. Liu, J., van Sommeren, S. & Huang, H. e. a. Association analyses identify 38 susceptibility loci for inflammatory bowel disease and highlight shared genetic risk across populations. *Nature Genetics* **47,** 979–986. `https://www.nature.com/articles/ng.3359` (2015) (cit. on p. 5).

74. *Downloadable inflammatory bowel disease predictor* `http://ftp.ebi.ac.uk/pub/databases/gwas/summary_statistics/GCST003001-GCST004000/GCST003043/IBD_trans_ethnic_association_summ_stats_b37.txt.gz`. Accessed: 2022-05-30 (cit. on p. 5).

75. Malik, R., Chauhan, G. & Traylor, M. e. a. Multiancestry genome-wide association study of 520,000 subjects identifies 32 loci associated with stroke and stroke subtypes. *Nature Genetics* **50,** 524–537. `https://www.nature.com/articles/s41588-018-0058-3` (2018) (cit. on p. 5).

76. *Downloadable stroke predictor* `http://ftp.ebi.ac.uk/pub/databases/gwas/summary_statistics/GCST006001-GCST007000/GCST006908/harmonised/29531354-GCST006908-HP_0002140.h.tsv.gz`. Accessed: 2022-05-30 (cit. on p. 5).

77. Wray, N., Ripke, S. & Mattheisen, M. e. a. Genome-wide association analyses identify 44 risk variants and refine the genetic architecture of major depression. *Nature Genetics* **50,** 668–681. https://www.nature.com/articles/s41588-018-0090-3 (2018) (cit. on p. 5).

78. *Downloadable major depressive disorder predictor* https://www.med.unc.edu/pgc/download-results/mdd/. Accessed: 2022-05-30 (cit. on p. 5).

79. Of the Psychiatric Genomics Consortium, S. W. G. Biological insights from 108 schizophrenia-associated genetic loci. *Nature* **511,** 421–427. https://www.nature.com/articles/nature13595 (2014) (cit. on p. 6).

80. *Downloadable schizophrenia predictor* https://www.med.unc.edu/pgc/download-results/scz/. Accessed: 2022-05-30 (cit. on p. 6).

81. Privé, F. *et al.* Portability of 245 polygenic scores when derived from the UK Biobank and applied to 9 ancestry groups from the same cohort. *American Journal of Human Genetics* **109,** 12–23. ISSN: 15376605 (2022) (cit. on p. 8).
